# Geographical variation in dementia: systematic review with meta-analysis

**DOI:** 10.1101/2023.12.11.23299178

**Authors:** K.E. Walesby, S.D. Shenkin, J.K. Burton, C. Dunlop, C. Fenton, D Gray, L.A. McGillicuddy, J.M. Starr, T. Wilkinson, G. Muniz Terrera, T.C. Russ

**Affiliations:** Alzheimer Scotland Dementia Research Centre and Centre for Cognitive Ageing and Cognitive Epidemiology, 7 George Square, University of Edinburgh, UK; Ageing and Health Research Group, Centre for Population Health Sciences, and Advanced Care Research Centre, Usher Institute, University of Edinburgh; Academic Geriatric Medicine, School of Cardiovascular and Metabolic Health, College of Medical, Veterinary & Life Sciences, University of Glasgow; Old Age Psychiatry, Edinburgh Royal Infirmary, NHS Lothian, UK; NESSIE, Usher Institute, University of Edinburgh; Fairnington Centre, Hexham General Hospital, Cumbria, Northumberland, Tyne and Wear NHS Foundation Trust (CNTW), UK; Old Age Psychiatry, Royal Edinburgh Hospital, NHS Lothian, UK; Centre for Clinical Brain Sciences, University of Edinburgh, UK; Department of Social Medicine, Ohio University, Ohio University Heritage College of Osteopathic Medicine and Edinburgh Dementia Prevention, University of Edinburgh, UK

**Keywords:** Dementia, Alzheimer disease, epidemiology, geography

## Abstract

**Background:** Understanding geographical variation of dementia could highlight important modifiable socio-environmental risk factors. A previous systematic review (2012) identified an increased risk of Alzheimer dementia in rural living in High-Income Countries (HICs), with a dearth of studies in Low to Middle-Income Countries (L-MICs). We updated this review to examine geographical variations in dementia, to encompass the growing number of studies in this field.

**Methods:** We systematically reviewed the literature for cross-sectional or longitudinal observational studies that compared dementia incidence or prevalence between two or more geographical areas including rural and urban settings.

We conducted a narrative synthesis of included papers. Where possible, we undertook meta-analysis, generating odds ratios for rural versus urban dementia prevalence and stratified the analysis by HICs and L-MICs.

**Results:** We identified 38 relevant papers, encompassing approximately 98,502,147 people. Twenty-seven papers were included in the quantitative synthesis. Study methodologies varied widely. Dementia rates varied geographically (0.43-38.5%). Overall, rural living was associated with small increased odds of dementia (OR, 1.20, 95% CI 1.03-1.40; *P* value = 0.0182). Stratification by HICs and L-MICs demonstrated further variation, with increased odds of dementia in rural areas in L-MICs but not HICs.

**Conclusions:** There is some evidence of geographical variation of dementia. Rural living was associated with small increased odds of dementia, with stratification showing evidence in rural areas of L-MICs but not HICs. We believe this has not been reported previously. Future research must consider life course geographical exposure and addressing heterogeneity in definitions of ‘rural’ and ‘urban.’

**What this study adds:** We confirm that rural living (compared to urban living) is associated with a small increased odds of dementia (OR 1.20, 95%CI 1.03-1.40). We demonstrate for the first time that this is driven by increased odds of dementia in rural areas in Low to Middle-Income Countries (L-MICs) rather than High Income Countries (HICs), and that the odds of dementia were higher in urban areas in large studies in HICs. Future studies need to carefully consider study setting, method of dementia ascertainment, when exposures may occur, and risk of bias, to understand the role of environment and geography in dementia risk.

## INTRODUCTION

Dementia is a growing worldwide public health concern. The epidemiology of this complex syndrome is not yet fully understood.^1^ Age and known genetic, health, and lifestyle risk factors play an important role in dementia incidence, but together they do not fully explain dementia aetiology. Life course geographical variation might influence dementia risk or protective factors.

A systematic review in 2012 found “evidence of geographical variation in rates of dementia in affluent countries at a variety of geographical scales.”^2^ Furthermore, an increased risk of Alzheimer’s dementia was associated with rural living (Prevalence OR 1.50, 90% CI 1.33-1.69), with early life rural living further increasing this risk to approximately double (OR 2.22, 90%CI 1.19-4.16). However, these results were based on very few studies, and there have been several subsequent studies. We therefore updated this systematic review^2^ to reflect the developing body of evidence in this field. In our present systematic review, we report a comprehensive overview of the literature specifically examining the influence of rural and urban living on risk of dementia.

## METHODS

### Registration and reporting

We prospectively registered the protocol with PROSPERO, reference CRD42016050323 (http://www.crd.york.ac.uk/) and report the review in accordance with the Preferred Reporting of Items in Systematic Reviews and Meta-Analyses (PRISMA) guidance.^3,4^

### Information sources

We searched multiple databases on 18^th^-31st October 2018 for papers since the published review^2^ search date of 15^th^ April 2010: search information and indicative search strategies were developed with an Information Specialist (CF), (Appendix 1).

### Eligibility criteria

Eligible studies included: cross-sectional or longitudinal studies of any duration comparing primary dementia rates (prevalence or incidence) between two or more different geographical sites. Our initial search was broad then focused to rural/urban; therefore, many studies (117) were excluded as ‘wrong comparator.’

We excluded studies purely on young onset dementia (<60 years old) or mild cognitive impairment. We did not restrict to English language (Appendix 2).

### Study selection and data collection

Two reviewers (from KEW, JKB, and TCR) independently screened all titles and abstracts using Covidence,^5^ and two then reviewed full text with conflicts resolved by discussion. Google translate was used for data extraction from papers not in English.

Two of KEW, LAM, CD, or DG extracted data using a bespoke and piloted form with discrepancies resolved by discussion and reference to the original paper.

Where articles did not report odds ratios for dementia risk in rural and urban areas,^6–16^ we computed these if the relevant raw data were reported or contacted authors^9,17–20^ : data provided by two authors.^18,19^ We contacted authors regarding full-text publications from abstracts identified, and overlap between studies. When both unadjusted and multivariable-adjusted odds ratios were reported, we report unadjusted odds ratio. When both DSM-IV and 10/66 method were used for dementia ascertainment, we report the 10/66 results^21–23^ (as this has shown greater dementia ascertainment in LICs and LMICs^24,25^) except when the 10/66 data were incomplete.^26^

### Strategy for data synthesis

Where sufficient data were available, we conducted a random effects meta-analysis of odds ratios. Post-hoc we decided to explore the data by stratifying by Higher Income Countries (HICs) and Low to Middle-Income Countries (L-MICs)^27^ (Appendix 3), and further by study size. We recognise this classification of country income can change with time (Appendix 3).

Between-study heterogeneity was assessed by the *I*^2^ statistic. Studies which could not be combined quantitatively were reported in a narrative synthesis.

### Risk of bias assessment in individual studies

KEW, SDS, and TCR assessed the risk of bias using an adapted Risk of Bias Assessment Tool for Non-Randomized studies (RoBANS), (Appendix 4), with disagreements resolved by discussion.

## RESULTS

The narrative synthesis includes 38 new papers (**Table 1),** 27 of which – including 33 cohorts - could be meta-analysed (**Figure 1**).

**Figure 1:**
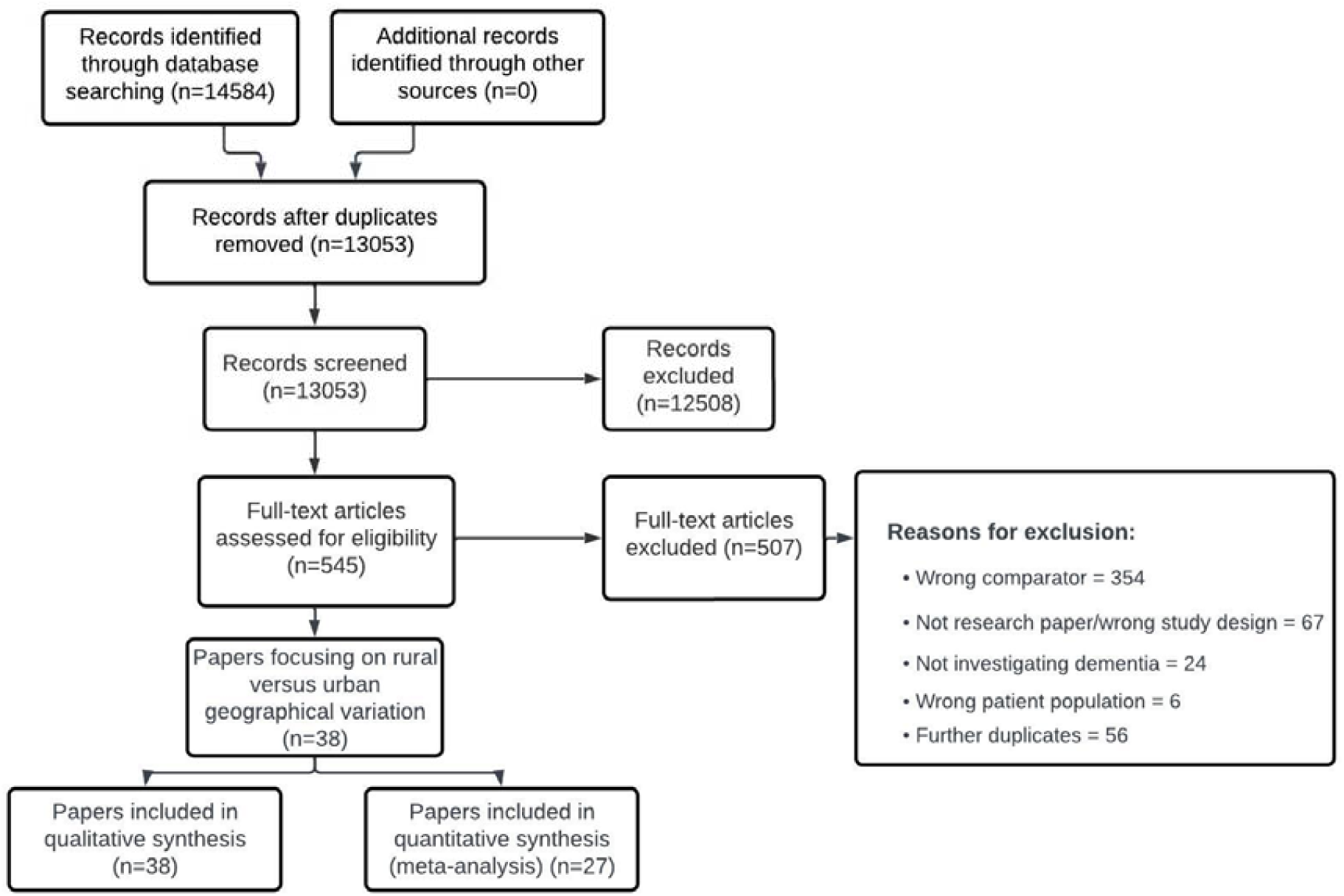
PRISMA flowchart detailing selection of studies for inclusion in the review: rural/urban differences in dementia.

**Table 1:**
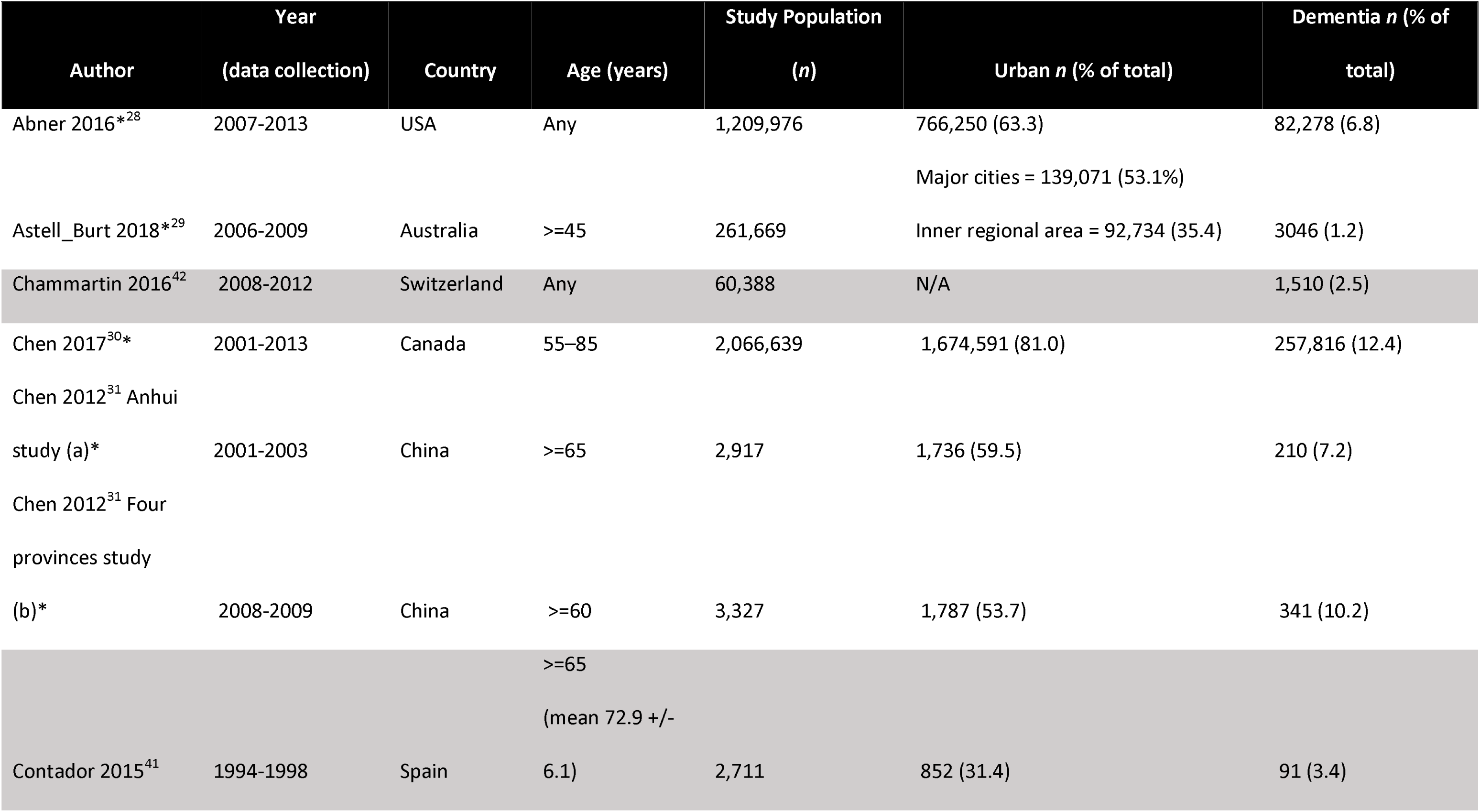
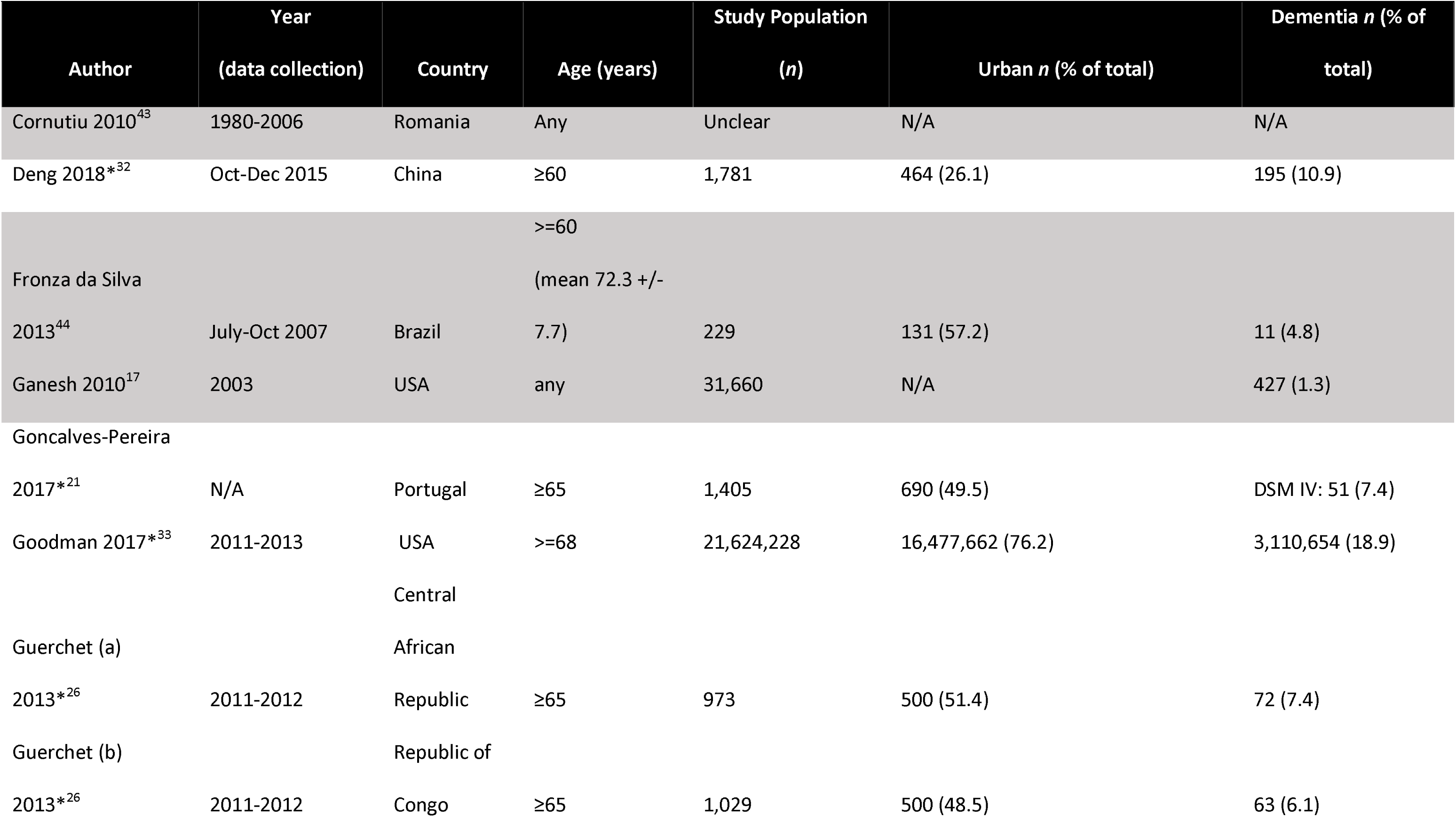
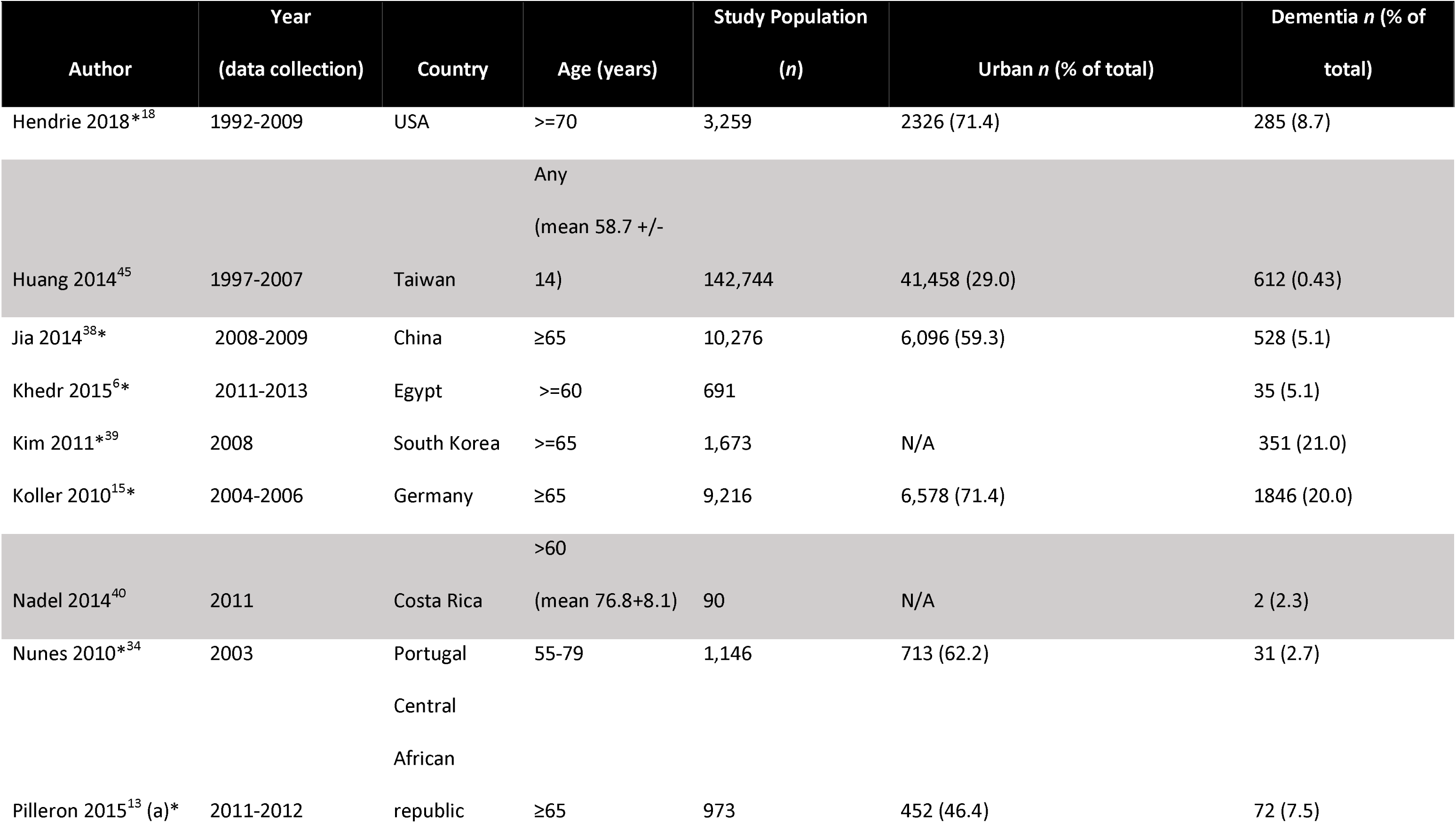
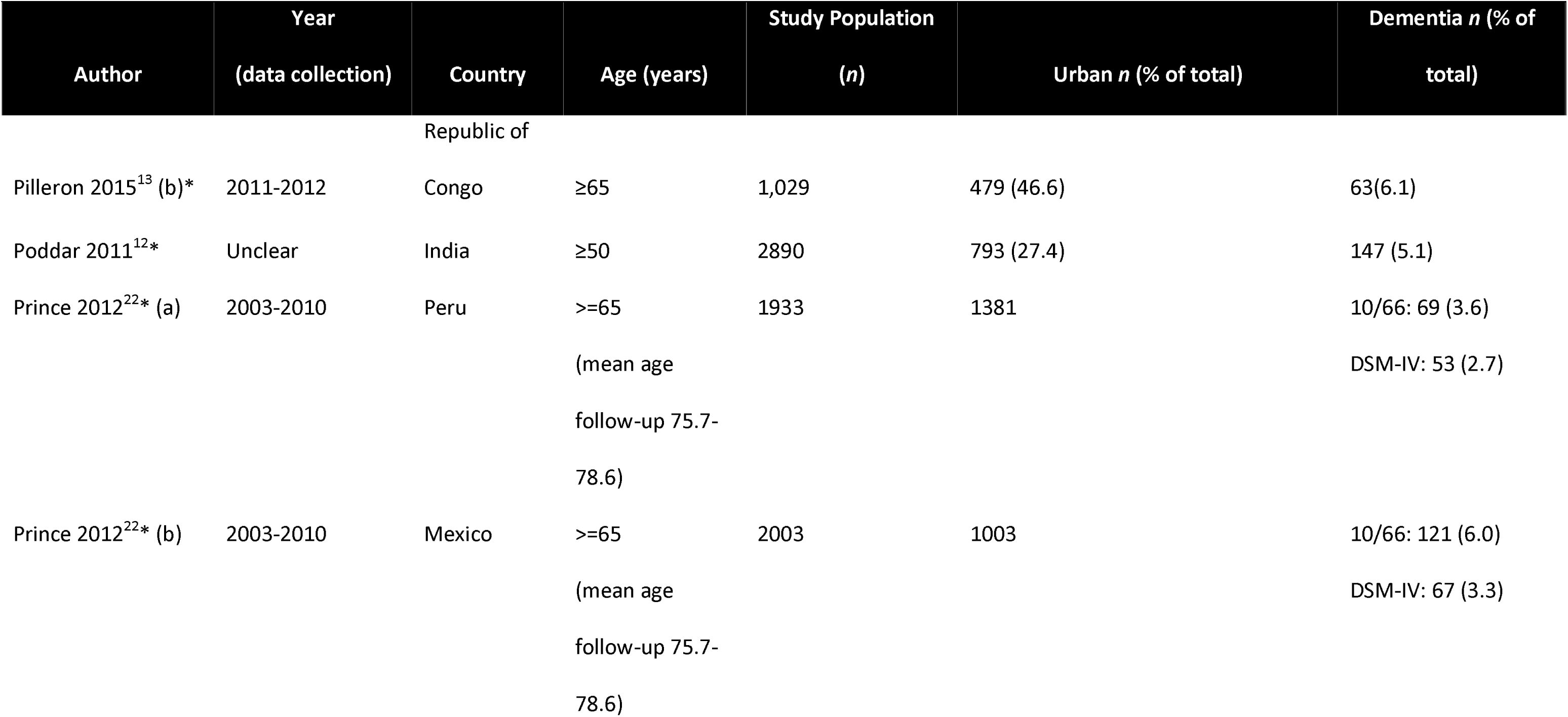
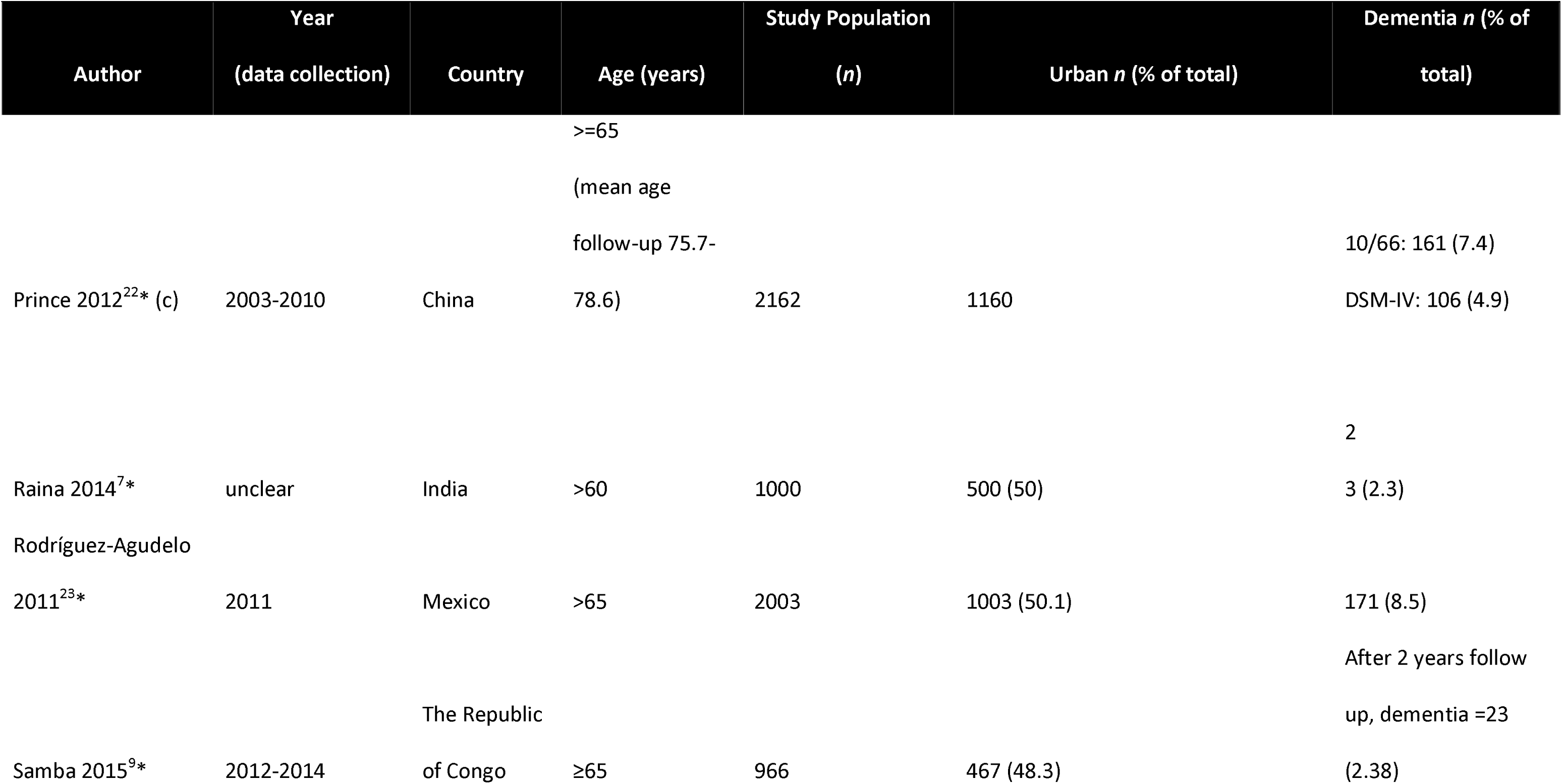
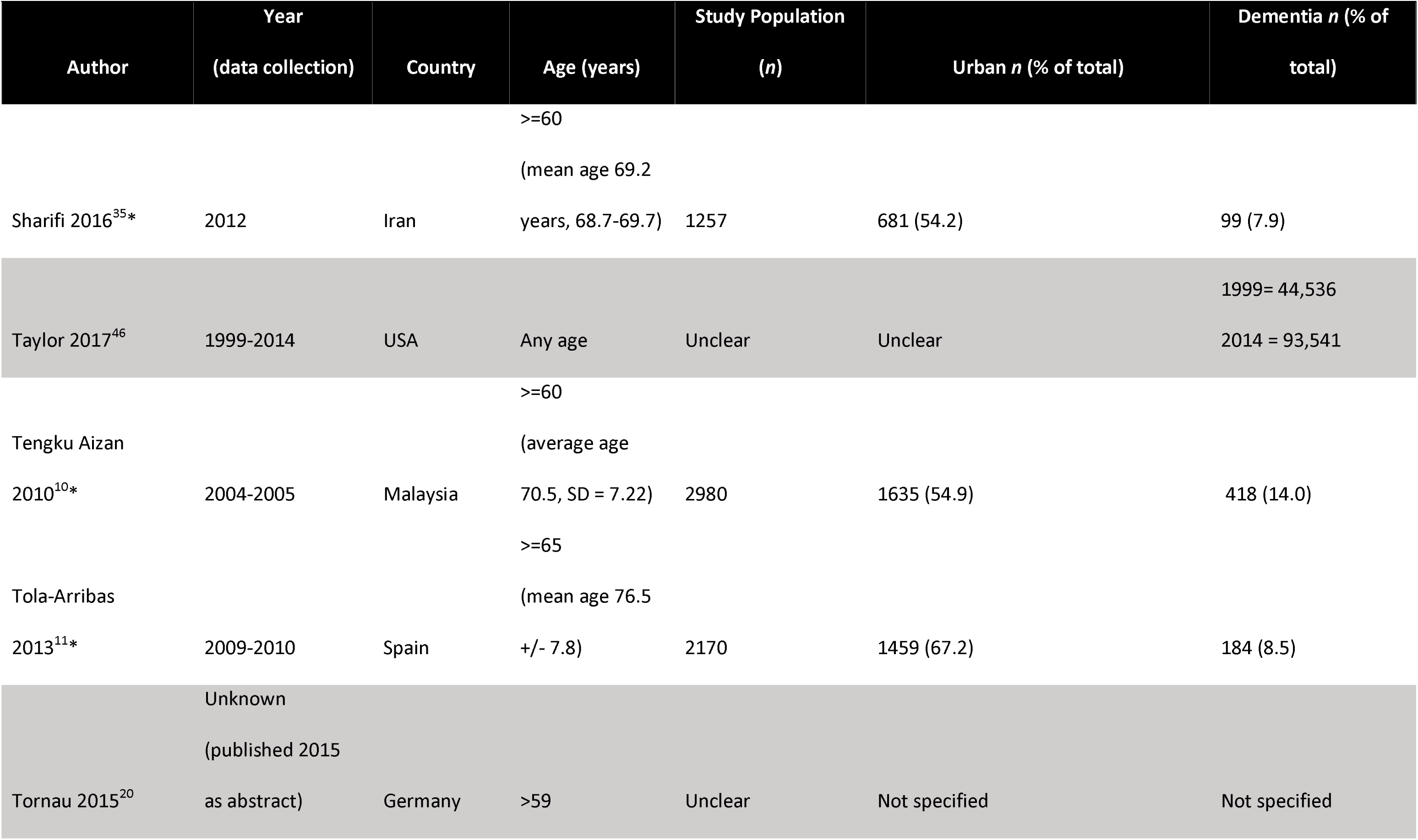
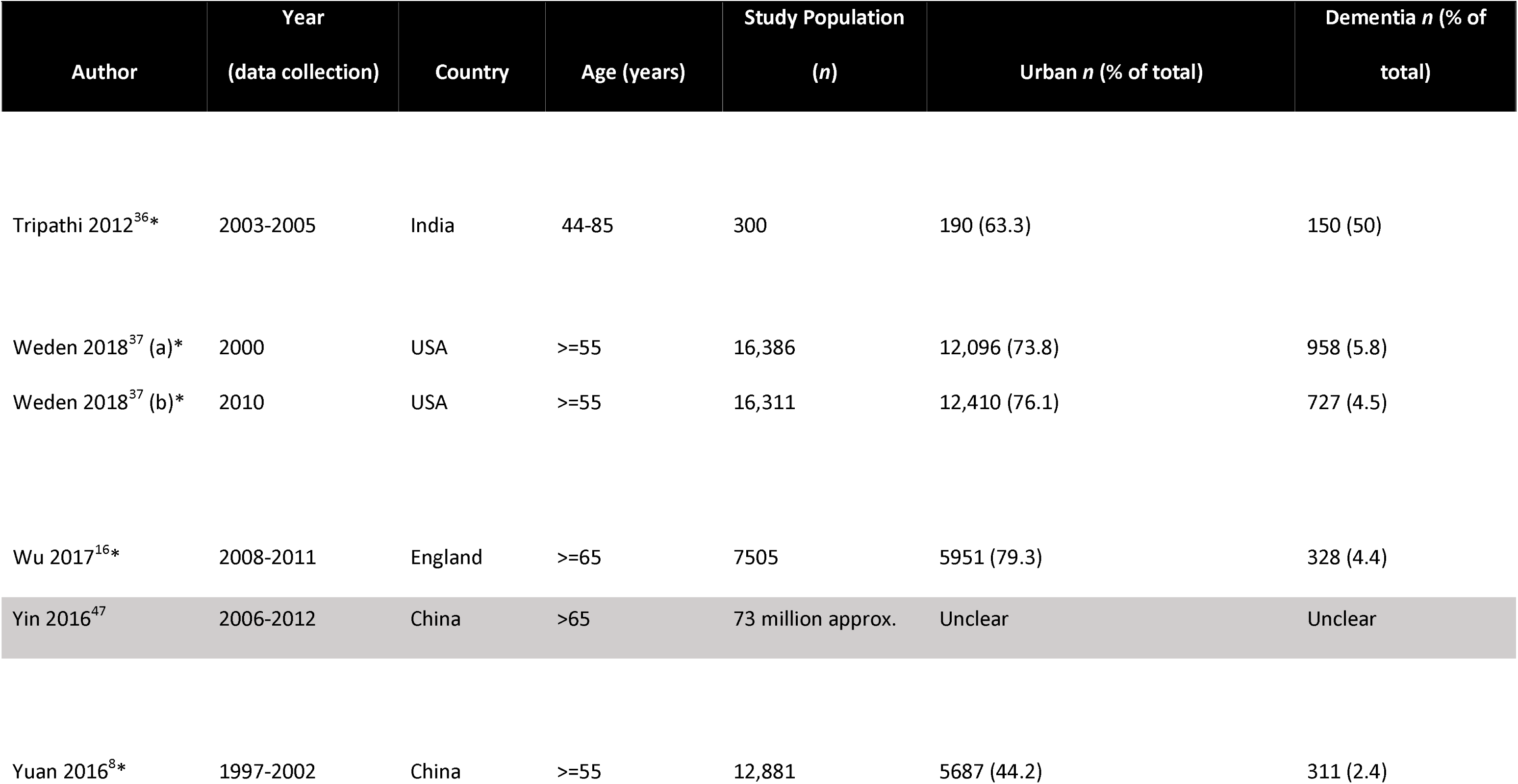
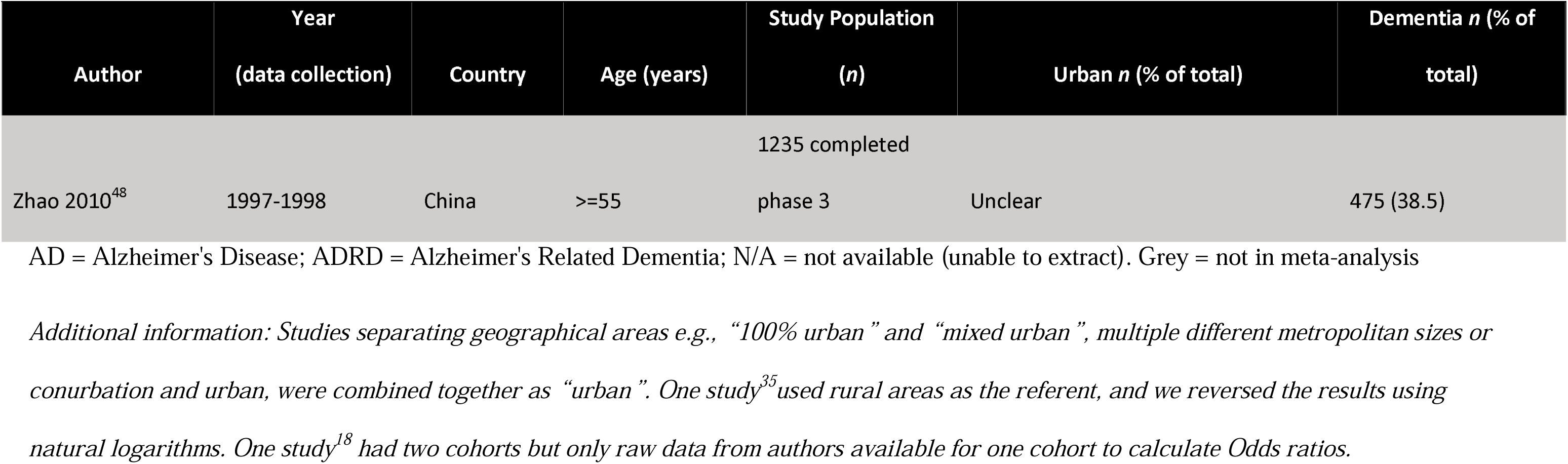
Executive summary of studies meeting inclusion criteria: rural/urban differences in dementia.

### Geographical locations and sample sizes

Studies were predominantly in the Americas (N=11) and Western Pacific region (especially China, N=11), and also Europe (N=9) South-East Asia (N=4), Africa (N=3), and Eastern Mediterranean region (N=2) (one study covered three areas). Of the meta-analysis papers, 12 papers (13 studies) were based in HICs and 15 papers (20 studies) in L-MICs.

In the 35 papers that included a sample size, this ranged from 90 to 73,000,000 people, median 2170. Of 27 papers in the meta-analysis, study samples ranged from 300 to 21,624,228 people, median 2162.^6–11,13,15,16,18,19,21,22,28–39^ The definition of ‘rural’ and ‘urban’ was heterogeneous. Too few papers included information on education, deprivation, or other socio-economic factors in relation to rural or urban settings and subsequent dementia risk to include in the meta-analysis. Three papers considered childhood and adulthood rural or urban residence and risk of dementia,^18,40,41^ only one was included in the meta-analysis.^18^

### Dementia diagnosis

There was considerable heterogeneity in dementia ascertainment methodologies (**Appendix 5, Supplementary Table 1),** mostly using surveys/interviews, clinical assessment and ICD codes. Dementia prevalence ranged from 0.43%-38.5% of the study populations in all studies, and 2.4-21% in the meta-analysis studies. Few identified dementia subtype. Most papers compared dementia against non-dementia or ‘normal’, with some including cognitive impairment/MCI in the ‘no dementia’ group, and others excluding those with MCI.

### Quantitative synthesis

Overall, rural living was associated with small increased odds of dementia, compared to urban living (OR, 1.20, 95% CI 1.03-1.40, *P* value *= 0.0182*; N=25,274,741, N_Dementia_=3,462,039; 33 studies), **Figure 2**. However, stratifying by HICs and L-MICs status revealed a difference, with HICs studies showing no effect of rural vs urban (1.01, 0.88 to 1.16, *P* value = 0.8842; N=25,221,600, N_Dementia_=3,458,633; 13 studies) and L-MICs suggesting increased odds of dementia in rural areas (1.33, 1.06 −1.68, *P* value = 0.0141; N=53,141, N_Dementia_=3406 20 studies). There was high heterogeneity: *I*^2^ value = 99.78% (for all studies), 99.72% (for only HICs studies) and 87.64% (for only L-MICs studies).

**Figure 2:**
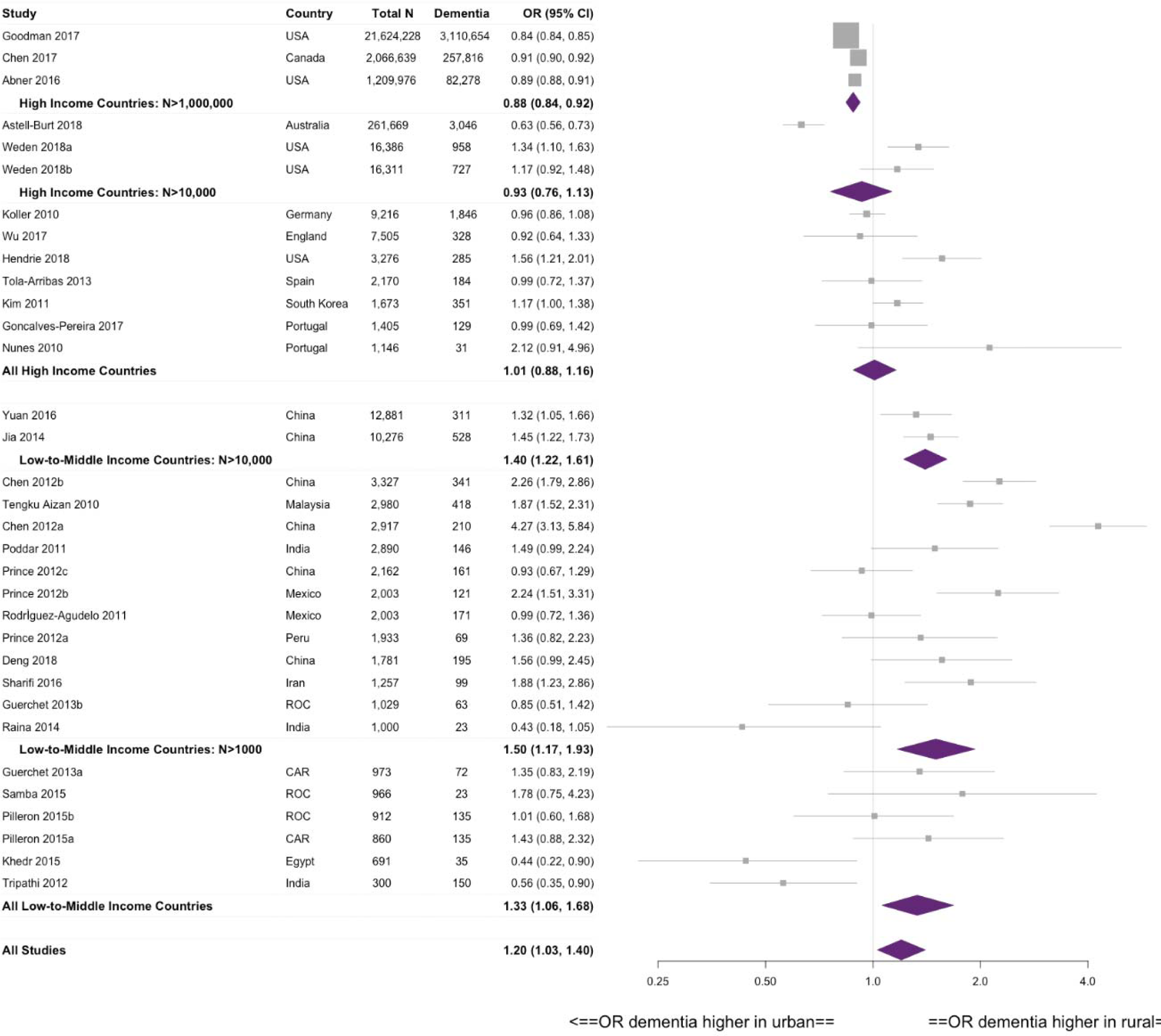
Forest plot showing meta-analysed rural/urban odds ratios of dementia stratified by HICs/L-MICs status and size of study: rural/urban differences in dementia. ^1^. ^1^Guerchet 2013, Pilleron 2015, Prince 2012 (Mexico only) Rodríguez-Agudelo 2011 and Samba 2015: unclear if there is overlap in these L-MICs papers (as more than one similar study in the same country). Sensitivity analysis removing the smaller cohorts (Pilleron 2015, Rodríguez-Agudelo 2011 and Samba 2015) shows no significant change in result

Sensitivity analysis to explore the impact of potential overlap in L-MICs study participants^9,13,22,23,26^ did not significantly affect the results: rural living was still linked with a slight increase in dementia risk (1.35, 1.02-1.79, *P* value = 0.0354, *I*^2^ = 90.69%). Including studies from the previous systematic review also showed a slight increased risk of dementia in rural areas, (1.18, 1.01-1.37) (**Appendix 6**).

Further stratifying HICs and L-MICs by study size confirmed in larger L-MICs studies (N>10,000) higher odds of dementia in rural areas (or lower in urban areas; 1.56, 1.20-2.02; N=23,157, N_Dementia_=839; 2 studies), unchanged when moderate-sized studies (N>1000) were included (1.50, 1.017-1.93; N=48,439, N_Dementia_=2856; 14 studies). However, larger HICs studies (N>1,000,000) showed *lower* odds of dementia in rural areas (0.88, 0.84-0.92; N=24,900,843, N_Dementia_=3,450,748; 3 studies). This effect was reduced when moderate-sized studies (N>10,000) were included (0.93, 0.76-1.13; N=25,195,209, N_Dementia_=3,455,479; six studies). Studies in L-MICs were generally smaller than those in HICs.

### Narrative synthesis of non-meta-analysis studies

The 11 heterogeneous papers not included in the meta-analysis, were undertaken in Europe (Switzerland, Romania, Germany and Spain),^20,41–43^ South-America (Brazil),^44^ Central America (Costa Rica),^40^ North America (USA)^17,46^ and Asia (Taiwan and China).^45,47,48^ between 1993 and 2012. One study investigated dementia in diabetic patients,^45^ the other studies investigated dementia in general. Five studies had no age restriction, ^17,42,43,45,46^ the others included people >55/59 years or >60/65 years.^20,40,41,44,47,48^, with sample size from 90 to approximately 73 million (median, 17,186).

The dementia ascertainment methodologies varied greatly: data linkage with ICD codes; ^17,20,42,43,45–47^ cross sectional incidence surveys; questionnaires; structured interviews and/or screening by clinical assessment.^40,41,44,48^

Just over half of these studies reflected the findings of the meta-analysis, with an increase of dementia in rural areas.^41,43,44,46–48^ The results were unclear in several of these studies,^20,40^ and showed no difference in one study investigating the variation of AD drug dispensing^17^. Of the two studies suggesting an increase in urban areas, one was specifically investigating dementia (AD) deaths, ^42^ and the other newly diagnosed diabetic patients.^45^

Most studies only assessed the relationship between residence at one time point and subsequent dementia, but one study^41^ investigated childhood and adulthood residence, finding childhood rural residence was linked with a non-significant trend for risk of dementia (HR = 1.52, 95% CI = 0.93-2.49, p=0.08), which became significant in adulthood (HR 1.61, 95% CI 1.06-2.46, p<0.05).^41^, but there was no association in late-life (HR 0.78, 95% CI 0.49-1.23, p=0.29).^41^

### Risk of Bias

**Figures 3** and **Appendix 7** show the risk of bias assessments for included studies. There was great variation in the risk of bias and, in particular, high risk in measurement of exposure (geographical location) in approximately half of the studies.

**Figure 3.**
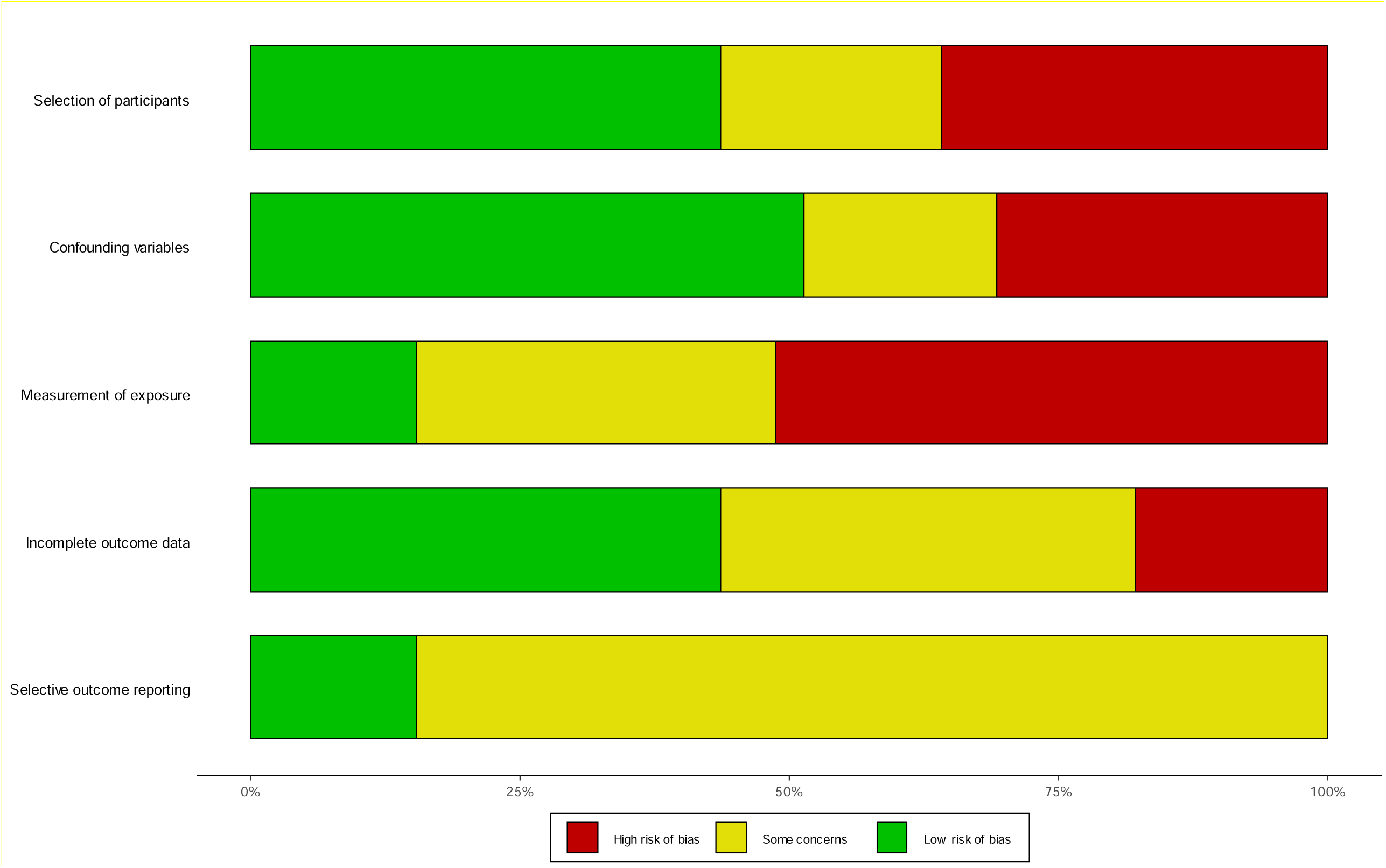
Summary of Risk of Bias assessment: rural/urban differences in dementia.

## DISCUSSION

Overall, rural living was associated with small increased odds of dementia compared to urban living (OR, 1.20, 95% CI 1.03-1.40, *P* value = 0.0182; N=25,274,741, N_Dementia_=3,462,039; 33 studies). Further stratification by HICs and L-MICs showed this difference was mostly driven by L-MICs (1.33, 1.06 −1.68, *P* value = 0.0141; N=53,141, N_Dementia_=3406 20 studies). In large HICs, odds of dementia were higher in urban areas (0.88, 0.84-0.92). To our knowledge, this difference has not been reported before, and has important implications for research and policy in this area. Studies were heterogeneous in the definition of ‘rural’ or ‘urban’ and identification of dementia, and there was high risk of bias in the exposure (geographical variation). Studies often did not report when in the life-course the geographical exposure was measured, and whether this changed across their lifetime, and the association between rurality and dementia prevalence and incidence may change with at different ages.

Increased risk of dementia in those living in rural areas may be due to specific aspects of living in these geographical locations, such as access to healthcare services/dementia diagnosis, air pollution or social connections, or may be confounded or mediated by education or socio-economic status.^1^ There is a complex interplay between the environment and the individual living in it. For dementia risk, this includes direct and indirect associations between the environment, and cognition, and noting that both of these can change across the life course.^49^

Dementia prevalence can vary country to country, influenced by the drive to seek a dementia diagnosis as well as the different the methodologies used for dementia ascertainment. A recent systematic review recommends using multiple sources, including access to medical records, to ascertain dementia to improve consistency in methodologies between studies.^50,51^

For example, in another systematic review^52^ which found a mixed picture between green space exposure and dementia risk, green space exposure was associated with higher incidence of dementia measured by anti-dementia medication and reversed if the other methods for detecting dementia were used.^53^ It is therefore essential that the method(s) of dementia ascertainment is clearly reported and the potential limitations and biases of this acknowledged.

When comparing multiple studies it is important that the dementia ascertainment rate for the study population is calculated and compared with an expected rate.^51^ Dementia diagnosis, particularly in L-MICs, can still be challenging, and there is ongoing debate about the best criteria for dementia diagnosis:^54^ criteria (such as the 10/66 criteria) may have greater accuracy than DSM-IV in low levels of literacy.^54^ Furthermore, in rural areas in general, or L-MICs, there may be limited access to diagnostic assessment, ^55^ especially if services are centralised.^56^ Our systematic review did not show an increased risk of dementia in urban settings to suggest this bias, but this needs considered when data are included from more countries.

There may be increased cognitive reserve in people living in urban areas ^49,57,58^, particularly during childhood, which influences dementia variation between rural and urban living. The impact from social deprivation may be higher than that from rural or urban location (e.g. the odds of a cognitive disorder were twice as high in those living in poor-quality rural environments, than those in rural areas).^16^

Education influences dementia risk, ^59^ but few studies investigated education in relation to the geographical area across time. Socio-economic status (SES) also influences dementia prevalence rates^50,60^. Our systematic review was unable to assess for SES in influencing the difference in rural versus urban dementia risk. Whilst education, SES and other variables for overall dementia risk were often reported, these variables were rarely considered in relation to rural or urban areas. Further work is required to understand the complex interplay between geographical location, education, socioeconomic circumstances, prior cognitive ability and dementia risk: whether there are critical time points in a person’s life where individual or combined factors influence dementia risk. Future studies should include more detailed information on participants’ education, SES measures in relation to geographical area, as well as the different definitions of geographical categorisation.

### Comparison with other literature

This large and comprehensive systematic review builds on a previous systematic review in 2010 which showed, in 25 rural versus urban studies, that there was some evidence of greater all-cause dementia odds with rural living [prevalence OR = 1.11, 90% CI 0.79-1.57; incidence OR = 1.20, 90% CI 0.84-1.71], and that the association between rurality and dementia risk was stronger in studies only including people with Alzheimer Disease (OR=1.50, 90% CI 1.33-1.69), but weaker in studies only including people with vascular dementia (OR= 1.09, 90% CI 0.65-1.83). The review also suggested that exposure to rural living early in life further increases the risk of subsequent dementia.

In comparison, our subsequent review which included 38 new papers was unable to investigate the dementia sub-type risk, or the life course effect of geographical location, as too few studies which reported adequate data on these predictors to undertake meaningful meta-analysis. Most studies used dementia as a ‘blanket’ term encompassing different sub-types of dementia, and did not provide adequate data for the life course effect of geographical location on dementia risk. The previous review investigated dementia prevalence and incidence at a number of geographical scales (country-to-country comparisons, rural to urban comparisons, and regional comparisons), but there has been a rapid increase studies published on this topic, therefore our current review focussed on rural to urban comparisons.

### Limitations and strengths: of the systematic review

The literature search for this comprehensive systematic review was developed with an information specialist and used a validated comprehensive search strategy from our previous review, with a pre-registered protocol, and was reported according to PRISMA guidance. At least two reviewers independently reviewed titles and abstracts, and extracted and analysed the studies with high levels of agreement using proforma which was produced and trialled for data extraction, and risk of bias (adapting the ROBANS tool). Authors were contacted for additional information, to increase data available for meta-analysis. The language was not limited to English to minimise bias in selecting papers for review.

The limitations of this study include that despite a comprehensive search strategy including forward citation of included studies; it is possible that relevant papers may have been missed, as we did not search for grey literature. There is also significant variation within and between countries of the level of geography which is recorded.

The Risk of Bias assessment tool was based on ROBANS and whilst it did assess the quality of the method of dementia ascertainment, for the studies using big data and ICD-10 codes it was not possible to assess the quality of that dementia diagnosis.

### Limitations and strengths: of the included studies

We identified 38 new relevant papers in the last 5 years, and were able to extract, or calculate, odds ratios for 27 papers (33 studies) for the meta-analysis, including studies from a range of L-MICs and HICs. The sample sizes were overall large (range 90 to 73,000,000, median 2170 for all papers, and 300 to 21,624,228, median 2162, for those in the meta-analysis) and the method of defining dementia was reported in the studies.

The individual studies were heterogeneous in their methodologies. In particular, there was significant heterogeneity in of the definition of ‘rural’ and ‘urban’, and how this translates at an international level for comparison. Too few studies reported the impact of geographical location at different time points in a person’s life to include in meta-analysis. This means there could be confounding or bias influencing the risk of dementia.

There is a need for standard definitions of urbanisation and rurality, and a clearer definition of “place.”^49^ Despite proposed quantifiable measures of urbanisation through population density and sprawl and further evaluation of physical characteristics of proximal environments,^49^ none of our included papers considered these factors.

It is also important to acknowledge that diagnostic coverage, those diagnosed with dementia rather than the true dementia prevalence, can be low^61^ and therefore missed cases (particularly within studies using large big datasets of routinely collected data^62^) may introduce bias, potentially under-reporting, in interpreting geographical variation of dementia versus traditional face-to-face epidemiological studies.

### Implications

There has been a rapid increase in interest in the geographical predictors of dementia, therefore this study, limited only to urban versus rural geographical variation, confirmed previous findings of increased odds of subsequent dementia for people living in rural areas. The heterogeneity of the studies means understanding rural versus urban risk of dementia is complicated. To ensure a future robust study to investigate the geographical variation of rural versus urban living and dementia risk (given our finding of OR of 1.16 to 1.26), the sample size would need to be approximately 1000-4 000 people.

Further consensus on the term ‘geographical location’ with clear definitions of ‘rural’ and ‘urban’ that is translatable across, and within, different countries is important to understand the complex interplay between the environment and the individual in relation to dementia risk. Future large-scale studies must also include detailed information about the area (including a measure of deprivation, greenspace, access to services etc.) and a person’s geographical location across time.

We know the incidence and prevalence of dementia is rising in L-MICs, but larger studies in this systematic review are in HICs, where they are more likely to have healthcare systems with routinely collected data allowing larger scale studies. Whereas, L-MICs are more likely to use traditional (smaller scale) face-to-face epidemiological studies. As these research methods are more costly than analysing nationally collected data, it results in fewer studies in L-MICs, with lower numbers of participants, but likely to provide more detailed analysis. Finally, the definition of LIC, HMIC and HIC has the potential to change with time, which is important to consider given the longitudinal aspect for dementia risk.

### Conclusions

Overall, rural living was associated with small-increased odds of dementia, compared to urban living (OR, 1.20, 95% CI 1.03-1.40, *P* value = 0.0182). Stratifying by HICs and L-MICs suggested increased odds of dementia in L-MICs rural areas, (1.33, 1.06 −1.68, *P* value = 0.0141). However, methodological differences complicate combining the extant literature. Therefore, better-designed studies with careful consideration of mediating and confounding variables are required to answer this important question definitively.

## Data Availability

All data produced in the present study are available upon reasonable request to the authors.

## Acknowledgements

KEW was supported by clinical research fellowship from Alzheimer Scotland and the University of Edinburgh Centre for Cognitive Ageing and Cognitive Epidemiology, part of the cross-council Lifelong Health and Wellbeing initiative (MR/L501530/1). Funding from the Biotechnology and Biological Sciences Research Council (BBSRC) and Medical Research Council (MRC) is gratefully acknowledged. KEW is also supported by Alzheimer Scotland Dementia Research Centre.

JKB is supported by an NHS Education for Scotland/Chief Scientists Office Postdoctoral Clinical Lectureship (PCL/21/01).

GMT is supported by the Osteopathic Heritage Foundation Ralph S. Licklider D.O.

Lastly, we would like to thank Professor John Starr (co-author) for his huge lifetime contribution to dementia research worldwide. We are deeply saddened by his sudden unexpected death on 9 December 2018 and miss our colleague greatly. His input as co-author was important to this research.

