## Appendix 1 for "Geographical variation in dementia: systematic review with meta-analysis"

**Appendix 1: Databases and search strategies**

**Databases searched: rural/urban differences in dementia**

| Database | Database start | Search date |
|---|---|---|
| ASSIA | 1987 | 18.10.18 |
| MEDLINE | 1950 | 18.10.18 |
| EMBASE | 1974 | 18.10.18 |
| CINAHL | 1981 | 18.10.18 |
| GeoRef | 1669 | 18.10.18 |
| Global Health | 1973 | 18.10.18 |
| LILACS | 1982 | 23.10.18 |
| PsycINFO | 1806 | 18.10.18 |
| COPAC | 1996 | 23.10.18 |
| SciELO | 1997 | 23.10.18 |
| Dissertations and Theses | 1861 | 31.10.18 |
| PapersFirst | 1993 | 23.10.18 |
| ProceedingsFirst | 1993 | 23.10.18 |

The following databases used in our previous systematic review on this topic are no longer active and therefore were not included: FRANCIS, GEOBASE, Australian Digital Theses Program, ADT, Theses Canada Portal, Conference Papers Index

**Indicative search strategies for Ovid Medline, EMBASE, and ASSIA: rural/urban differences in dementia**

**Epub Ahead of Print, In-Process & Other Non-Indexed Citations, Ovid MEDLINE(R) Daily and Ovid MEDLINE(R)** 1946 to Present

| | |
|---|---|
| 1 | Dementia/ep |
| 2 | Alzheimer Disease/ep |
| 3 | Delirium/ep |
| 4 | Wernicke Encephalopathy/ep |
| 5 | Delirium, Dementia, Amnestic, Cognitive Disorders/ep |
| 6 | dement*.ti,ab. |
| 7 | alzheimer*.ti,ab. |
| 8 | (lewy* adj2 bod*).ti,ab. |
| 9 | deliri*.ti,ab. |
| 10 | (chronic adj2 cerebrovascular).ti,ab. |
| 11 | "organic brain disease".ti,ab. |

| | |
|---|---|
| 12 | "organic brain syndrome".ti,ab. |
| 13 | ("normal pressure hydrocephalus" and "shunt*").ti,ab. |
| 14 | "benign senescent forgetfulness".ti,ab. |
| 15 | (cerebr* adj2 deteriorat*).ti,ab. |
| 16 | (cerebr* adj2 deteriorat*).ti,ab. |
| 17 | (pick* adj2 disease).ti,ab. |
| 18 | huntington*.ti,ab. |
| 19 | binswanger*.ti,ab. |
| 20 | korsako*.ti,ab. |
| 21 | or/1-20 |
| 22 | Incidence/ |
| 23 | Prevalence/ |
| 24 | Epidemiologic Studies/ |
| 25 | Incidence.ti,ab. |
| 26 | Prevalence.ti,ab. |
| 27 | distribut*.ti,ab. |
| 28 | epidem*.ti,ab. |
| 29 | "geographical variation*".ti,ab. |
| 30 | or/22-29 |
| 31 | Geography/ |
| 32 | North America/ep |
| 33 | Pacific States/ep |
| 34 | Asia/ep |
| 35 | Africa/ep |
| 36 | Canada/ep |
| 37 | Europe/ep |
| 38 | Far East/ep |
| 39 | Scotland/ep |
| 40 | United States/ep |
| 41 | Sweden/ep |
| 42 | Spain/ep |
| 43 | (Geograph* or cluster* or AFGHANISTAN or "ÅLAND ISLANDS" or ALBANIA or SAMOA or ANDORRA or ANGOLA or ANGUILLA or ANTARCTICA or ANTIGUA or BARBUDA or ARGENTINA or ARMENIA or ARUBA or AUSTRALIA or AZERBAIJAN or BAHAMAS or BANGLADESH or BARBADOS or BELGIUM or BELIZE or BERMUDA or BOLIVIA or BONAIRE or BOSNIA).tw. |
| 44 | (HERZEGOVINA or BOTSWANA or "BOUVET ISLAND" or BRAZIL or "BRITISH INDIAN OCEAN TERRITORY" or BRUNEI or DARUSSALAM or BULGARIA or "BURKINA FASO" or BURUNDI or CAMBODIA or CAMEROON or CANADA or CAPE VERDE or "CAYMAN ISLANDS" or "CENTRAL AFRICAN REPUBLIC" or CHAD or CHILE or CHINA or "CHRISTMAS ISLAND").tw. |
| 45 | ("COCOS ISLANDS" or "KEELING islands" or COLOMBIA or COMOROS or CONGO or "COOK ISLANDS" or "COSTA CÔTE" or CROATIA or CUBA or |

| | |
|---|---|
| | CURACAO or CYPRUS or CZECH* or DENMARK or DJIBOUTI or DOMINICA or "DOMINICAN REPUBLIC" or ECUADOR or EGYPT or "EL SALVADOR").tw. |
| 46 | ("EQUATORIAL GUINEA" or ERITREA or ESTONIA or ETHIOPIA or "FALKLAND ISLANDS" or "FAROE ISLANDS" or FIJI or FINLAND or FRANCE or GUIANA or POLYNESIA or "FRENCH SOUTHERN TERRITORIES" or GABON or GAMBIA or GEORGIA or GERMANY or GHANA or GIBRALTAR or GREECE).tw. |
| 47 | (GREENLAND or GRENADA or GUADELOUPE or GUAM or GUATEMALA or GUERNSEY or GUINEA or "GUINEA-BISSAU" or HAITI or "HOLY SEE" or HONDURAS or "HONG KONG" or HUNGARY or ICELAND or INDIA or INDONESIA or IRAN or IRAQ or IRELAND or "ISLE OF MAN" or ISRAEL or ITALY or JAMAICA or JAPAN or JERSEY or JORDAN or KAZAKHSTAN or KENYA).tw. |
| 48 | (KIRIBATI or KOREA or KUWAIT or KYRGYZSTAN or LAO or LATVIA or LEBANON or LESOTHO or LIBERIA or LIBYA or LIECHTENSTEIN or LITHUANIA or LUXEMBOURG or MACAO or "MO MAC" or MACEDONIA or MADAGASCAR or MALAWI or MALAYSIA or MALDIVES or MALI or MALTA).tw. |
| 49 | ("MARSHALL ISLANDS" or MARTINIQUE or MAURITANIA or MAYOTTE or MEXICO or MICRONESIA or MOLDOVA or MONACO or MONGOLIA or MONTSERRAT or MOROCCO or MOZAMBIQUE or MYANMAR or NAMIBIA or NAURU or NEPAL or NETHERLANDS or "NEW CALEDONIA" or "NEW ZEALAND").tw. |
| 50 | (NICARAGUA or NIGER or NIGERIA or NIUE or "NORFOLK ISLAND" or "NORTHERN MARIANA ISLANDS" or NORWAY or OMAN or PAKISTAN or PALAU or PALESTINE or PANAMA or "PAPUA NEW GUINEA" or PARAGUAY or PERU or PHILIPPINES or PITCAIRN or POLAND or PORTUGAL or PUERTO RICO or QATAR or REUNION or ROMANIA or "RUSSIAN FEDERATION").tw. |
| 51 | (RWANDA or "SAINT BARTHÉLEMY" or "SAINT HELENA" or ASCENSION or "TRISTAN DA CUNHA" or "SAINT KITTS" or "SAINT LUCIA" or "SAINT MARTIN" or "SAINT PIERRE" or "SAINT VINCENT" or SAMOA or "SAN MARINO" or "SAO TOME" or "SAUDI ARABIA" or SENEGAL or SERBIA or SEYCHELLES or "SIERRA LEONE" or SINGAPORE or "SINT MAARTEN").tw. |
| 52 | (SLOVAKIA or SLOVENIA or "SOLOMON ISLANDS" or SOMALIA or "SOUTH AFRICA" or "SOUTH GEORGIA" or SUDAN or SPAIN or "SRI LANKA" or SUDAN or SURINAME or SVALBARD or SWAZILAND or SWEDEN or SWITZERLAND or "SYRIAN ARAB REPUBLIC" or TAIWAN or TAJIKISTAN or TANZANIA or THAILAND or TIMOR-LESTE or TOGO or TOKELAU or TONGA).tw. |
| 53 | (TRINIDAD or TOBAGO or TUNISIA or TURKEY or TURKMENISTAN or "TURKS AND CAICOS ISLANDS" or TUVALU or UGANDA or UKRAINE or "UNITED ARAB EMIRATES" or "UNITED KINGDOM" or "UNITED STATES" or "URUGUAY" or UZBEKISTAN or VANUATU or VENEZUELA or "VIET NAM" or "VIRGIN ISLANDS" or "WALLIS AND FUTUNA").tw. |
| 54 | ("WESTERN SAHARA" or YEMEN or ZAMBIA or ZIMBABWE or Great Britain or England or Scotland or Wales or America* or Syria or RUSSIA or Vietnam).tw. |
| 55 | or/31-54 |

| | |
|---|---|
| 56 | 21 AND 30 AND 55 |

**Embase** 1947-Present, updated daily via OVID

| | |
|---|---|
| 1 | exp dementia/ep [Epidemiology] |
| 2 | exp Alzheimer disease/ep [Epidemiology] |
| 3 | exp delirium/ep [Epidemiology] |
| 4 | exp cognitive defect/ep [Epidemiology] |
| 5 | exp Wernicke encephalopathy/ep [Epidemiology] |
| 6 | dement*.ti,ab. |
| 7 | alzheimer*.ti,ab. |
| 8 | (lewy* adj2 bod*).ti,ab. |
| 9 | deliri*.ti,ab. |
| 10 | (chronic adj2 cerebrovascular).ti,ab. |
| 11 | "organic brain disease".ti,ab. |
| 12 | "organic brain syndrome".ti,ab. |
| 13 | ("normal pressure hydrocephalus" and "shunt*").ti,ab. |
| 14 | "benign senescent forgetfulness".ti,ab. |
| 15 | (cerebr* adj2 deteriorat*).ti,ab. |
| 16 | (cerebr* adj2 insufficient*).ti,ab. |
| 17 | (pick* adj2 disease).ti,ab. |
| 18 | huntington*.ti,ab. |
| 19 | binswanger*.ti,ab. |
| 20 | korsako*.ti,ab. |
| 21 | or/1-20 |
| 22 | exp incidence/ |
| 23 | exp prevalence/ |
| 24 | Epidemiologic Studies/ |
| 25 | Incidence.ti,ab. |
| 26 | Prevalence.ti,ab. |
| 27 | distribut*.ti,ab. |
| 28 | epidem*.ti,ab. |
| 29 | "geographical variation*".ti,ab. |
| 30 | or/22-29 |
| 31 | 21 AND 30 |
| 32 | (Geograph* or cluster* or AFGHANISTAN or "ÅLAND ISLANDS" or ALBANIA or SAMOA or ANDORRA or ANGOLA or ANGUILLA or ANTARCTICA or ANTIGUA or BARBUDA or ARGENTINA or ARMENIA or ARUBA or AUSTRALIA or AZERBAIJAN or BAHAMAS or BANGLADESH or BARBADOS or BELGIUM or BELIZE or BERMUDA or BOLIVIA or BONAIRE or BOSNIA).tw. |
| 33 | (HERZEGOVINA or BOTSWANA or "BOUVET ISLAND" or BRAZIL or "BRITISH INDIAN OCEAN TERRITORY" or BRUNEI or DARUSSALAM or BULGARIA or "BURKINA FASO" or BURUNDI or CAMBODIA or CAMEROON or CANADA or CAPE VERDE or "CAYMAN ISLANDS" or "CENTRAL AFRICAN REPUBLIC" or CHAD or CHILE or CHINA or "CHRISTMAS ISLAND").tw. |
| 34 | ("COCOS ISLANDS" or "KEELING islands" or COLOMBIA or COMOROS or CONGO or "COOK ISLANDS" or "COSTA CÔTE" or CROATIA or CUBA or |

| | |
|---|---|
| | CURACAO or CYPRUS or CZECH* or DENMARK or DJIBOUTI or DOMINICA or "DOMINICAN REPUBLIC" or ECUADOR or EGYPT or "EL SALVADOR").tw. |
| 35 | ("EQUATORIAL GUINEA" or ERITREA or ESTONIA or ETHIOPIA or "FALKLAND ISLANDS" or "FAROE ISLANDS" or FIJI or FINLAND or FRANCE or GUIANA or POLYNESIA or "FRENCH SOUTHERN TERRITORIES" or GABON or GAMBIA or GEORGIA or GERMANY or GHANA or GIBRALTAR or GREECE).tw. |
| 36 | (GREENLAND or GRENADA or GUADELOUPE or GUAM or GUATEMALA or GUERNSEY or GUINEA or "GUINEA-BISSAU" or HAITI or "HOLY SEE" or HONDURAS or HONG KONG or HUNGARY or ICELAND or INDIA or INDONESIA or IRAN or IRAQ or IRELAND or "ISLE OF MAN" or ISRAEL or ITALY or JAMAICA or JAPAN or JERSEY or JORDAN or KAZAKHSTAN or KENYA).tw. |
| 37 | (KIRIBATI or KOREA or KUWAIT or KYRGYZSTAN or LAO or LATVIA or LEBANON or LESOTHO or LIBERIA or LIBYA or LIECHTENSTEIN or LITHUANIA or LUXEMBOURG or MACAO or "MO MAC" or MACEDONIA or MADAGASCAR or MALAWI or MALAYSIA or MALDIVES or MALI or MALTA).tw. |
| 38 | ("MARSHALL ISLANDS" or MARTINIQUE or MAURITANIA or MAYOTTE or MEXICO or MICRONESIA or MOLDOVA or MONACO or MONGOLIA or MONTSERRAT or MOROCCO or MOZAMBIQUE or MYANMAR or NAMIBIA or NAURU or NEPAL or NETHERLANDS or "NEW CALEDONIA" or "NEW ZEALAND").tw. |
| 39 | (NICARAGUA or NIGER or NIGERIA or NIUE or "NORFOLK ISLAND" or "NORTHERN MARIANA ISLANDS" or NORWAY or OMAN or PAKISTAN or PALAU or PALESTINE or PANAMA or "PAPUA NEW GUINEA" or PARAGUAY or PERU or PHILIPPINES or PITCAIRN or POLAND or PORTUGAL or PUERTO RICO or QATAR or REUNION or ROMANIA or "RUSSIAN FEDERATION").tw. |
| 40 | (RWANDA or "SAINT BARTHÉLEMY" or "SAINT HELENA" or ASCENSION or "TRISTAN DA CUNHA" or "SAINT KITTS" or "SAINT LUCIA" or "SAINT MARTIN" or "SAINT PIERRE" or "SAINT VINCENT" or SAMOA or "SAN MARINO" or "SAO TOME" or "SAUDI ARABIA" or SENEGAL or SERBIA or SEYCHELLES or "SIERRA LEONE" or SINGAPORE or "SINT MAARTEN").tw. |
| 41 | (SLOVAKIA or SLOVENIA or "SOLOMON ISLANDS" or SOMALIA or "SOUTH AFRICA" or "SOUTH GEORGIA" or SUDAN or SPAIN or "SRI LANKA" or SUDAN or SURINAME or SVALBARD or SWAZILAND or SWEDEN or SWITZERLAND or "SYRIAN ARAB REPUBLIC" or TAIWAN or TAJIKISTAN or TANZANIA or THAILAND or TIMOR-LESTE or TOGO or TOKELAU or TONGA).tw. |
| 42 | (TRINIDAD or TOBAGO or TUNISIA or TURKEY or TURKMENISTAN or "TURKS AND CAICOS ISLANDS" or TUVALU or UGANDA or UKRAINE or "UNITED ARAB EMIRATES" or "UNITED KINGDOM" or "UNITED STATES" or "URUGUAY" or UZBEKISTAN or VANUATU or VENEZUELA or "VIET NAM" or "VIRGIN ISLANDS" or "WALLIS AND FUTUNA").tw. |
| 43 | ("WESTERN SAHARA" or YEMEN or ZAMBIA or ZIMBABWE or Great Britain or England or Scotland or Wales or America* or Syria or RUSSIA or Vietnam).tw. |
| 44 | OR/32-43 |
| 45 | 31 AND 44 |

**Applied Social Sciences Index & Abstracts (ASSIA)** via Proquest
(ti(Dementia) OR ab(Dementia) OR ti("Alzheimer Disease") OR ab("Alzheimer Disease") OR ti(Delirium) OR ab(Delirium) OR ti("Wernicke Encephalopathy") OR ab("Wernicke Encephalopathy") OR ti("Cognitive Disorder*") OR ab("Cognitive Disorder*") OR ti(huntington*) OR ab(huntington*) OR ti(Aphasia) OR ab(Aphasia) OR ti(Creutzfeldt-Jakob) OR ab(Creutzfeldt-Jakob) OR ti(CADASIL) OR ab(CADASIL) OR ti(Lewy Body) OR ab(Lewy Body) OR ti(Pick Disease) OR ab(Pick Disease)) AND (ti(Prevalence) OR ab(Prevalence) OR ti(incidence) OR ab(incidence) OR ti(distribut*) OR ab(distribut*) OR ti(epidem*) OR ab(epidem*)) AND (ti(Geograph*) OR ab(Geograph*) OR ti(cluster*) OR ab(cluster*) OR ti(Afghanistan) ab(Afghanistan) OR ti(Albania) OR ab(Albania) OR ti(Algeria) OR ab(Algeria) OR ti(Angola) OR ab(Angola) OR ti(Antarctica) OR ab(Antarctica) OR ti(Argentina) OR ab(Argentina) OR ti(Armenia) OR ab(Armenia) OR ti(Australia) OR ab(Australia) OR ti(Austria) OR ab(Austria) OR ti(Azerbaijan) OR ab(Azerbaijan) OR ti(Bahamas) OR ab(Bahamas) OR ti(Bahrain) OR ab(Bahrain) OR ti(Bangladesh) OR ab(Bangladesh) OR ti(Barbados) OR ab(Barbados) OR ti(Belarus) OR ti(Belgium) OR ab(Belgium) OR ti(Belize) OR ab(Belize) OR ti(Benin) OR ab(Benin) OR ti(Bermuda) OR ab(Bermuda) OR ti(Bolivia) OR ab(Bolivia) OR ti(Bosnia) OR ab(Bosnia) OR ti(Botswana) OR ab(Botswana) OR ti(Brazil) OR ab(Brazil) OR ti(Brunei) OR ab(Brunei) OR ti(Bulgaria) OR ab(Bulgaria) OR ti(Burkina fasso) OR ab(Burkina fasso) OR ti(Burma) OR ab(Burma) OR ti(Burundi) OR ab(Burundi) OR ti(Cambodia) OR ab(Cambodia) OR ti(Cameroon) OR ab(Cameroon) OR ti(Canada) OR ab(Canada) OR ti(Central African republic) or ab(Central African republic) OR ti(Chad) OR ab(Chad) OR ti(Chile) OR ab(Chile) OR ti(China) OR ab(China) OR ti(Colombia) OR ab(Colombia) OR ti(Congo) OR ab(Congo) OR ti(Costa Rica) OR ab(Costa Rica) Or ti(Cote divoire) OR ab(Cote divoire) OR ti(Croatia) OR ab(Croatia) OR ti(Cuba) OR ab(Cuba) OR ti(Cyprus) OR ab(Cyprus) OR ti(Czech republic) OR ab(Czech republic) OR ti(Denmark) OR ab(Denmark) OR ti(Dominica) OR ab(Dominica) OR ti(Dominican republic) OR ab(Dominican republic) OR ti(Ecuador) OR ab(Ecuador) OR ti(Egypt) OR ab(Egypt) OR ti(El Salvador) OR ab(El Salvador) OR ti(England) OR ab(England) OR ti(Equatorial guinea) OR ab(Equatorial guinea) OR ti(Eritrea) OR ab(Eritrea) OR ti(Estonia) OR ab(Estonia) OR ti(Ethiopia) OR ab(Ethiopia) OR ti(Finland) OR ab(Finland) OR ti(France) OR ab(France) OR ti(Georgia) OR ab(Georgia) OR ti(Germany) OR ab(Germany) OR ti(Ghana) OR ab(Ghana) OR ti(Greece) OR ab(Greece) OR ti(Greenland) OR ab(Greenland) OR ti(Guinea) OR ab(Guinea) OR ti(Hong kong) OR ab(Hong kong) OR ti(Hungary) OR ab(Hungary) OR ti(Iceland) OR ab(Iceland) OR ti(India) OR ab(India) OR ti(Indonesia) OR ab(Indonesia) OR ti(Iran) OR ab(Iran) OR ti(Iraq) OR ab(Iraq) OR ti(Ireland) OR ab(Ireland) OR ti(Israel) OR ab(Israel) OR ti(Italy) OR ab(Italy) OR ti(Jamaica) OR ab(Jamaica) OR ti(Japan) OR ab(Japan) OR ti(Jordan) OR ab(Jordan) OR ti(Kazakhstan) OR ab(Kazakhstan) OR ti(Kenya) OR ab(Kenya) OR ti(Korea) OR ab(Korea) OR ti(Kuwait) OR ab (Kuwait) OR ti(Kyrgyzstan) OR ab(Kyrgyzstan) OR ti(Laos) OR ab(Laos) OR ti(Latvia) OR ab(Latvia) OR ti(Lebanon) OR ab(Lebanon) OR ti(Lesotho) OR ab(Lesotho) OR ti(Libya) OR ab(Libya) OR ti(Lithuania) OR ab(Lithuania) OR ti(Malawi) OR ab(Malawi) OR ti(Malaysia) OR ab(Malaysia) OR ti(Mali) OR ab(Mali) OR ti(Mexico) OR ab(Mexico) OR ti(Mongolia) OR ab(Mongolia) OR ti(Morocco) OR ab(Morocco) OR ti(Mozambique) OR ab(Mozambique) OR ti(Namibia) OR ab(Namibia) OR ti(Nepal) OR ab(Nepal) OR ti(Netherlands) OR ab(Netherlands) OR ti(New Zealand) OR ab(New Zealand) OR ti(Nicaragua) OR ab(Nicaragua) OR ti(Niger) OR ab(Niger) OR ti(Nigeria) OR ab(Nigeria) OR ti(Norway) OR ab(Norway) OR ti(Pakistan) OR ab(Pakistan) OR ti(Papua new Guinea) OR ab(Papua new Guinea) OR ti(Paraguay) OR ab(Paraguay) OR ti(Peru) OR ab(Peru) OR ti(Philippines) OR ab(Philippines) OR ti(Poland) OR ab(Poland) OR ti(Portugal) OR

ab(Portugal) OR ti(Puerto Rico) OR ab(Puerto Rico) OR ti(Romania) OR ab(Romania) OR ti(Russia) OR ab(Russia) OR ti(Rwanda) OR ab(Rwanda) OR ti(Saudi Arabia) OR ab(Saudi Arabia) OR ti(Scotland) OR ab(Scotland) Or ti(Senegal) OR ab(Senegal) OR ti(Serbia) OR ab(Serbia) OR ti(Sierra Leone) OR ab(Sierra Leone) OR ti(Singapore) OR ab(Singapore) OR ti(Slovakia) OR ab(Slovakia) OR ti (Slovenia) OR ab(Slovenia) OR ti(Somalia) OR ab(Somalia) OR ti(South Africa) OR ab(South Africa) OR ti(Spain) Or ab(Spain) OR ti(Sri lanka) OR ab(Sri lanka) OR ti(Sudan) OR ab(Sudan) OR ti(Swaziland) OR ab(Swaziland) OR ti(Sweden) OR ab(Sweden) OR ti(Switzerland) OR ab(Switzerland) OR ti(Syria) OR ab(Syria) OR ti(Taiwan) OR ab(Taiwan) OR ti(Tajikistan) OR ab(Tajikistan) OR ti(Tanzania) OR ab(Tanzania) OR ti(Thailand) OR ab(Thailand) OR ti(Togo) OR ab(Togo) OR ti(Trinidad) OR ab(Trinidad) OR ti(Tunisia) OR ab(Tunisia) OR ti(Tunisia) OR ab(Tunisia) OR ti(Turkey) OR ab(Turkey) OR ti(Turkmenistan) OR ab(Turkmenistan) OR ti(Uganda) OR ab(Uganda) OR ti(Ukraine) OR ab(Ukraine) OR ti(United Arab Emirates) OR ab(United Arab Emirates) OR ti(United kingdom) OR ab(United kingdom) OR ti(United states) OR ab(United states) OR ti(Uruguay) OR ab(Uruguay) OR ti(Uzbekistan) OR ab(Uzbekistan) OR ti(Venezuela) OR ab(Venezuela) OR ti(Vietnam) OR ab(Vietnam) OR ti(Wales) OR ab(Wales) OR ti(Zambia) OR ab(Zambia) OR ti(Zimbabwe) OR ab(Zimbabwe))
