## Appendix 2 for "Geographical variation in dementia: systematic review with meta-analysis"

**Eligibility criteria**

**Included:**

- Cross-sectional or longitudinal studies of any duration comparing primary dementia rates (prevalence or incidence) between two or more different geographical sites, at any geographical scale but including a comparison of rural and urban areas

**Excluded:**

- Studies were no comparison between two or more different geographical sites or if the comparison between groups did not use the same methodology
- Studies where dementia was a secondary feature of a chronic disease (alcohol or traumatic brain injury), Parkinson's disease, Huntington's disease and Creutzfeldt-Jakob disease.
- Studies purely on young onset dementia (<60 years old) or mild cognitive impairment. We did not restrict to English language.
