## Appendix 3 for "Geographical variation in dementia: systematic review with meta-analysis"

### Table of country classification based on income and different years/classification

| | Wellcome research classification of LIC and LMIC countries based on The Organisation for Economic Co-operation and Development (OECD)* | World Bank Definition 2018 Countries | World Bank Definition 2022-2023 Countries |
|---|---|---|---|
| **High Income Country (HIC)** | USA, UK, Canada, Australia, Switzerland, Spain, Portugal, Germany, Romania, South Korea, Taiwan, | USA, UK, Canada, Australia, Swizterland, Spain, Portugal, Germany, South Korea, Taiwan | USA, UK, Canada, Australia, Swizterland, Spain, Portugal, Germany, Romania, South Korea, Taiwan |
| **Upper Middle-Income Country (UMIC)** | | Romania, Peru, Mexico, Brazil, Costa Rica, China, Malaysia, Iran | Peru, Mexico, Brazil, Costa Rica, China, Malaysia |

| | | | |
|---|---|---|---|
| **Lower Middle-Income Country (LMIC)** | China, Central African Republic (CAR), Republic of Congo (ROC), | Republic of Congo (ROC) | India, Iran, Republic of Congo (ROC), Egypt |
| **Low Income Country (LIC)** | Costa Rica, Mexico, India, Peru, Brazil, Iran, Egypt, Malaysia, | India, Central African Republic (CAR), Egypt | Central African Republic (CAR) |

[1]

[1] **We used this definition for this systematic review.  For the meta-analysis, we compared all in the High Income Countries (HIC) as these were largely consistent across the years with all other Income countries (Upper Middle-Income, Lower Middle-Income and Low Income Countries) and referred to this group as Low-to-Middle Income Countries (L-MIC)*
