## Appendix 4 for "Geographical variation in dementia: systematic review with meta-analysis"

**Appendix 4: Quality Assessment – Adapted ROBANs**

| Risk of Bias Item | Grading | Rationale/Examples |
|---|---|---|
| **Selection bias** | Low | Sampling frame clearly described with reasonable inclusion/exclusion criteria provided, ideally allowing for inclusion of all participants within the geographical area being studied |
| | High | Exclusions based on availability of data (e.g.);studies only considering certain postcodes; exclusion criteria likely to reduce the representativeness of a possible area (e.g. only looking at comparing ethnic groups in one area and not taking into consideration place of birth); only including population if already part of another study; only including population if clinical or in certain clinical area (e.g. comparing General practitioner localities, only comparing catchment areas for hospitals, only comparing geographical areas with large number of nursing homes). |
| | Unclear | Sampling frame unclear, criteria for inclusion/exclusion not provided or explained |
| **Confounding variables** | Low | Multivariate model accounting for likely possible confounding variables |
| | High | No consideration of confounding variables; univariate analyses only<br>(e.g. areas with non-varied socioeconomic status that would influence risk factor for dementia separately to |

| | | |
|---|---|---|
| | | geographical factors; age; vascular risk factors; large number of GP surgeries or hospitals in vicinity; large number of nursing homes in vicinity). |
| | Unclear | Methods for analysis not clearly described or reported |
| **Measurement of exposure** *(performance bias)* | Low | Clearly described method on how data were collected and extracted<br>Best practice includes description of who performed data extraction, case definitions/descriptions of eligible conditions<br>(e.g. at point during the person's life was the geography measured; how it was measured; who collected the information; what detail was collected – not just 'town'). |
| | High | Missing data on key predictor variables |
| | Unclear | Methods for assessing predictor variables not clearly described |
| **Incomplete outcome data** *(attrition bias)* | Low | Outcomes assessed for all included participants (including how many included had cognition assessed/obtained and how many it was missing for) and what method (e.g. MMSE, electronic data with coding for dementia; other cognitive assessments; more than one cognitive assessment) |
| | High | Missing outcome assessments (e.g. no data on how many did not have cognition assessed/recorded; no data on how cognition was assessed/obtained) |

| | | |
|---|---|---|
| | Unclear | Outcome assessment reported as percentages without absolute values being presented, preventing assessment of completeness of outcome reporting |
| **Selective outcome reporting** *(reporting bias)* | Low | Reporting as per published protocol |
| | High | Evidence that reporting deviates from publically accessible protocol |
| | Unclear | No protocol publically available |
