## Appendix 5 for "Geographical variation in dementia: systematic review with meta-analysis"

**Appendix 5: Supplementary Table 1 – Additional information on methodologies and range of diagnostic tools used**

| Author | Methodology |
| --- | --- |
| Abner 2016*[1] | Dementia in Electronic health records. |
| Astell_Burt 2018*[2] | Data linkage of records for cholinesterase inhibitor prescriptions. |
| Chammartin 2016[3] | Dementia from death certificates ICD codes G30-G309 |
| Chen 2017[4]* | Ontario Population Health and Environment Cohort (ONPHEC) Incidence study. Dementia from hospital admission/physician claims/prescription (ICD codes). |
| Chen 2012[5] Anhui study (a)* | Survey/interviews. Dementia defined using the GMS and the Automated Geriatric Examination for Computer Assisted Taxonomy (AGECAT). |
| Chen 2012[5] Four provinces study (b)* | Survey/interviews. Dementia defined using the GMS and the Automated Geriatric Examination for Computer Assisted Taxonomy (AGECAT). |
| Contador 2015[6] | Two cross sectional incidence surveys, (1994-1995,1997-1998). Screened with WHO protocol for dementia and confirmed by clinical assessment. Childhood/adulthood residence and dementia risk investigated. |
| Cornutiu 2010[7] | Incidence study, data from annual hospitalizations for AD. NInCDS_ADRDA criteria. The study population was unclear as possibly it included 600 participants but also documented 246 in 2002. |
| Deng 2018*[8] | Questionnaire/interview & clinical assessment. MMSE, IADL, GDS. |

| | |
|---|---|
| Fronza da Silva 2013[9] | Study investigated pesticide use in rural and urban areas and dementia. Questionnaire by clinician. Carer completed if participant could not. |
| Ganesh 2010[10] | Thesis. Medicare Current Beneficiary Survey (MCBS) ICD-9-CM codes: claims or self-report for AD. |
| Goncalves-Pereira 2017*[11] | Cross-sectional survey. Dementia using 10/66 Dementia Research Group or DSM IV criteria. 10/66 data used for meta-analysis. |
| Goodman 2017*[12] | Prevalence study from Centers for Medicare & Medicaid (CMS) claims data (ICD-9-CM) |
| Guerchet (a) 2013*[13] | Interview/survey study in EPIDEMCA population: Community Screening Interview for Dementia (CSI-D), the GMS-AGECAT and the CERAD's 10-word list. DSM IV criteria used. 10/66 data not complete. Additional information from author allowing inclusion in meta-analysis. |
| Guerchet (b) 2013*[13] | Interview/survey study in EPIDEMCA population: Community Screening Interview for Dementia (CSI-D), the GMS-AGECAT and the CERAD's 10-word list. DSM IV criteria used. 10/66 data not complete. Additional information from author allowing inclusion in meta-analysis. |
| Hendrie 2018*[14] | Indianapolis-Ibadan project (IIDP) incidence study. Only Indianapolis people included in this study. Two cohorts (1992 and 2001). Investigated childhood/adulthood residence and dementia risk. Clinical evaluation of dementia using neuropsychological tests, standardized neurologic and physical examination and functional |

| | |
|---|---|
| | status review and structured interview from informant. Medical records also reviewed. |
| Huang 2014[15] | Newly diagnosed patients with diabetes (n= 71,433) and matched controls (n= 71,311). Electronic health records using Taiwan's National Health Insurance Research Database (AD claims (ICD-9-CM code: 331.0). We used 'Metropolis' in their definitions to extract the data for Urban location. |
| Jia 2014[16]* | China Cognition and Ageing Study (China Coast) cohort: cross-sectional survey/clinical assessment. Clinical Dementia Rating (CDR) score |
| Khedr 2015[17]* | Cross-sectional community survey & clinical assessment. DSM-IV diagnostic criteria. |
| Kim 2011*[18] | Population survey (MMSE-KC) then clinical evaluation. |
| Koller 2010[19]* | Diagnosis in Clinical records |
| Nadel 2014[20] | Survey. MMSE & structured interview. 'Severe cognitive impairment' appeared to be synonymous with dementia so this information was extracted for the dementia outcome. |
| Nunes 2010*[21] | Survey & clinical assessment. |
| Pilleron 2015[22] (a)* | Interview/survey study in EPIDEMCA population: DSM IV criteria. Cognitive testing using Community Screening Interview for Dementia (CSI-D). Informant (relative) to assess daily activities. Further subsequent clinical assessment. Further psychometric tests (Free and Cued Selective Reminding Test, Zazzo's cancellation task and Issac's Set Test of Verbal Fluency). |

| | |
|---|---|
| Pilleron 2015[22] (b)* | Interview/survey study in EPIDEMCA population: DSM IV criteria. Cognitive testing using Community Screening Interview for Dementia (CSI-D). Informant (relative) to assess daily activities. Further subsequent clinical assessment. Further psychometric tests (Free and Cued Selective Reminding Test, Zazzo's cancellation task and Issac's Set Test of Verbal Fluency). |
| Poddar 2011[23]* | Cross-sectional. Two phase- phase 1 interview/survey and phase 2 - Hindi Mental State Examination |
| Prince 2012[24]* (a) | The countries that had both urban and rural totals were Mexico, Peru and China.<br>This is the data for the Peru study.<br>10/66 Dementia Research Group population-based studies; interview. Community Screening Instrument for Dementia (CSI'D') COGSCORE. 2003-2007 one-phase population based surveys and then 2007-2010 for the incidence wave and follow up. Both 10/66 and DSM-IV criteria were used for establishing cases of dementia. |
| Prince 2012[24]* (b) | The countries that had both urban and rural totals were Mexico, Peru and China. This is the data for the Mexico study.<br>10/66 Dementia Research Group population-based studies; interview. Community Screening Instrument for Dementia (CSI'D') COGSCORE. 2003-2007 one-phase population-based surveys and then 2007-2010 for the incidence wave and follow up. Both 10/66 and DSM-IV criteria were used for establishing cases of dementia. |
| Prince 2012[24]* (c | The countries that had both urban and rural totals were Mexico, Peru and China. |

| | |
|---|---|
| | This is the data for the China study. 10/66 Dementia Research Group population-based studies; interview. Community Screening Instrument for Dementia (CSI'D') COGSCORE. 2003-2007 one-phase population-based surveys and then 2007-2010 for the incidence wave and follow up. Both 10/66 and DSM-IV criteria were used for establishing cases of dementia. |
| Raina 2014[25]* | No cohort was used. Two-phase process (screening and clinical phase). (Hindi Mental State Examination (HMSE) and modified version for tribal group, clinical assessment). Study also investigated tribal and migrant populations but excluded for the meta-analysis and only rural and urban included. |
| Rodríguez-Agudelo 2011[26]* | Part of the 10/66 Research Group studies. Investigating neuropsychiatric symptoms in those with and without dementia. Two areas selected. Initial eligibility interviews at home followed by further interviews at health centres. Group 10/66 detection algorithm used for dementia diagnosis. |
| Samba 2015[27]* | EPIDEMCA (Epidemiology of Dementia in Central Africa) - FU (Follow-up) study cohort. Longitudinal population-based incidence study (966 participants at baseline without dementia). Participants traced with Psychometrical tests and health and vascular events investigated since the last visit. Verbal autopsies performed when older person was deceased and a reliable information available. DSM-IV and NINCDS-ADRDA criteria used for dementia/Alzheimer disease diagnoses. |

| | |
|---|---|
| Sharifi 2016[28]* | Two-phase screening cross-sectional study with multistage cluster random sampling national survey (The National Elderly Health Survey, NEHS). Physical examination and interview. Dementia diagnosis in two phases: Brief Cognitive Assessment Tool (BCAT) from Iran and diagnosis of dementia by GPs based on DSM IV criteria. Those with severe depression were removed to ensure no 'pseudo-dementia'. |
| Taylor 2017[29] | The study population was unclear as this study investigated AD mortality for different years, but it was unclear what the overall population (denominator) was for those years. State-level and county-level death certificate date from the National Vital Statistics System were analysed analysing ICD 10 codes. |
| Tengku Aizan 2010[30]* | The mental Health and Quality of Life of Older Malaysians cohort. Multi-stage sampling door-to-door survey with face-to-face interviews. Geriatric Mental State (GMS) B3 version assessed mental health status and entered into AGECAT software programme. AGECAT-GMS diagnostic algorithm was used for dementia cases. Rural and urban total dementia cases were reported separately. For the purpose of this systematic review, we totalled both of these for the total dementia *n* in this table. |
| Tola-Arribas 2013[31]* | DEMINVALL cohort (cross-sectional, two-phase, door-to-door, population-based study from social security healthcare holders' registry). Phase 1: Screening tool (Spanish version 7-minute Screen Neurocognitive Battery (7MS) l, or Spanish version IQCODE, or Spanish version Kawas Dementia Questionnaire if deceased). Phase |

| | |
|---|---|
| | 2: clinical assessment [including Spanish version of Cambridge Examination of Mental Disorders of the Elderly and Clinical Dementia Rating (CDR)] |
| Tornau 2015[32] | The study population was unclear as the database was stated as having >20,000 patients. Billing data of all statutory health insurances (90% of population) from an urban and a rural region in Germany. ICD 10-GM codes |
| Tripathi 2012[33]* | Case control study in outpatient tertiary clinic (150 consecutive patients attending cognitive disorders clinic and 150 age and sex matched control subjects attending the outpatient as other patients' relatives). Folstein's Mini Mental Status Examination and clinical assessment (neurologist and neuropsychologist) including AIIMS neuropsychological battery (validated in Indian population) and MRI brain. |
| Weden 2018[34] (a)* | Two cohorts within the one paper – 2000 and 2010. This is the 2000 cohort. Health and Retirement Study (HRS) multi-panel cohort panel survey linked to Census assessments of urbanicity. Cognitive functioning measured using validated methodology or if self-reporting using 27-point modified Telephone Interview for Cognitive Status. A proxy respondent used for those unable/unwilling to answer. |
| Weden 2018[34] (b)* | Two cohorts within the one paper – 2000 and 2010. This is the 2010 cohort. Health and Retirement Study (HRS) multi-panel cohort panel survey linked to Census assessments of urbanicity. Cognitive functioning measured using validated methodology or if self- |

| | |
|---|---|
| | reporting using 27-point modified Telephone Interview for Cognitive Status. A proxy respondent used for those unable/unwilling to answer. |
| Wu 2017[35]* | Baseline interviews were collected between 2008-2011. Urban was defined as urban city and town and also conurbation. For the purpose of this systematic review, urban city and town and conurbation were totalled to give the Urban *n= 5951*. Cohort study, Cognitive Function and Ageing Study II (CFAS II). Secondary data analysis CFAS II population-based epidemiologic study - baseline interviews, primary care registration. Geriatric Mental Status, MMSE, and Automatic Geriatric Examination for Computer-Assisted Taxonomy organicity level >=3. |
| Yin 2016[36] | Not a cohort. Mortality study using ICD codes from China Mortality Surveillance System Disease Surveillance Point (DSP). The total study participants were unclear. |
| Yuan 2016[37]* | Baseline participation was 1997 with follow up in 1999 and then 2000-2002. Cohort study [subsample of the 1997 study Dementia subtypes in China: Prevalence in Beijing (northeast), Xi'an (northwest), Shanghai (southeast), and Chengdu, (southwest)]. Incidence study (three phase). Phase 1 (dementia screening), door-to-door screening survey using the Chinese version of MMSE (C-MMSE) and Chinese version of ADL (CADL). Phase II (clinical diagnostic workup) clinical interview/examination, then re-screening, then MDT discussion. The total study population *n* in this table were the total traced at the second follow-up period (2000- |

| | |
|---|---|
| | 2002). The Urban *n* listed in this table comprised the total traced at the second follow-up period (2000-2002). |
| Zhao 2010[38] | Three phases: screening, clinical diagnostic ascertainment (clinical assessment and battery of neuropsychological tests - including ADL, POD, FOM, VFT, BD, DS, HAMD, Hachinski Ischemia Scale, C-MMSE, GDS), clinical confirmation (repeated screening and diagnostic examinations at 6 months in phase II participants). |

AD = Alzheimer's Disease; ADRD = Alzheimer's Related Dementia; N/A = not available (unable to extract). Grey = not in meta-analysis

**Supplementary Table 1** shows the range of diagnostic tools used. Few identified dementia subtype. Most papers used a broad dementia label as their outcome, but some did investigate the specific subtype, such as Alzheimer or vascular dementia. The most used diagnostic tools for dementia ascertainment were surveys/interviews, clinical assessment and ICD codes.

However, for some they separated the groups into those with dementia, those with cognitive impairment/MCI and those without either. Other papers did not specifically separate MCI from dementia; therefore, some within the 'no dementia' group may have MCI. For the purpose of this study, we used those with probable dementia/dementia as the outcome and in one paper 'severe cognitive impairment' appeared synonymous with dementia so this was used.

1. Abner EL, Jicha GA, Christian WJ, Schreurs BG. Rural-urban differences in Alzheimer's disease and related disorders diagnostic prevalence in Kentucky and West Virginia. *Journal of Rural Health* 2016;**32**(3):314-320.
2. Astell-Burt T, Feng X. Is the risk of developing Alzheimer's disease really higher in rural areas? A multilevel longitudinal study of 261,669 Australians aged 45 years and older tracked over 11 years. *Health and Place* 2018;**54**:132-137.
3. Chammartin F, Probst-Hensch N, Utzinger J, Vounatsou P. Mortality atlas of the main causes of death in Switzerland, 2008-2012. *Swiss Medical Weekly* 2016;**146**:w14280.
4. Chen H, Kwong JC, Copes R, et al. Exposure to ambient air pollution and the incidence of dementia: A population-based cohort study. *Environment International* 2017;**108**:271-277.
5. Chen R, Ma Y, Wilson K, et al. A multicentre community-based study of dementia cases and subcases in older people in China--the GMS-AGECAT prevalence and socio-economic correlates. *International Journal of Geriatric Psychiatry* 2012;**27**(7):692-702.
6. Contador I, Bermejo-Pareja F, Puertas-Martin V, Benito-Leon J. Childhood and Adulthood Rural Residence Increases the Risk of Dementia: NEDICES Study. *Current Alzheimer Research* 2015;**12**(4):350-7.
7. Cornutiu G. The incidence and prevalence of Alzheimer's disease. *Neurodegenerative Diseases* 2010;**8**(1-2):9-14.
8. Deng J, Cao C, Jiang Y, et al. Prevalence and effect factors of dementia among the community elderly in Chongqing, China. *Psychogeriatrics* 2018;**18**(5):412-420.
9. Silva EFd, Paniz VMV, Laste G, Torres ILdS. Prevalência de morbidades e sintomas em idosos: um estudo comparativo entre zonas rural e urbana. *Ciência & Saúde Coletiva* 2013;**18**(4):1029-1040.
10. Ganesh C. The diffusion of prescription drugs for Alzheimer's disease among medicare beneficiaries [Ph.D.]. The Pennsylvania State University, 2010.
11. Goncalves-Pereira M, Cardoso A, Verdelho A, et al. The prevalence of dementia in a Portuguese community sample: a 10/66 Dementia Research Group study. *BMC Geriatrics* 2017;**17**(1):261.
12. Goodman RA, Lochner KA, Thambisetty M, et al. Prevalence of dementia subtypes in United States Medicare fee-for-service beneficiaries, 2011-2013. *Alzheimer's & Dementia* 2017;**13**(1):28-37.
13. Guerchet M, Ndamba-Bandzouzi B, Mbelesso P, et al. Comparison of rural and urban dementia prevalences in two countries of central africa: The epidemca study. *Alzheimer's and Dementia* 2013;**1)**:P688.
14. Hendrie HC, Smith-Gamble V, Lane KA, et al. The Association of Early Life Factors and Declining Incidence Rates of Dementia in an Elderly Population of African Americans. *Journals of Gerontology Series B: Psychological Sciences & Social Sciences* 2018;**73**:S82-S89.
15. Huang C, Chung C, Leu H, et al. Diabetes mellitus and the risk of Alzheimer's disease: a nationwide population-based study. *PLoS ONE* 2014;**9**(1).
16. Jia J, Wang F, Wei C, et al. The prevalence of dementia in urban and rural areas of China. *Alzheimer's & Dementia: The Journal of the Alzheimer's Association* 2014;**10**(1):1-9.
17. Khedr E, Fawi G, Abbas MA, et al. Prevalence of mild cognitive impairment and dementia among the elderly population of Qena Governorate, Upper Egypt: a community-based study. *Journal of Alzheimer's Disease* 2015;**45**(1):117-26.
18. Kim K, Park J, Kim M, et al. A nationwide survey on the prevalence of dementia and mild cognitive impairment in South Korea. *JAD, Journal of Alzheimer's Disease* 2011;**23**(2):281-291.
19. Koller D, Eisele M, Kaduszkiewicz H, et al. Ambulatory health services utilization in patients with dementia - is there an urban-rural difference? *International Journal of Health Geographics* 2010;**9**(59).

20. Nadel JL, Ulate D. Incidence and risk factors for cognitive impairment in rural elderly populations in Costa Rica^ien

Incidencia y factores de riesgo para la discapacidad cognitiva en poblaciones rurales de tercera edad en Costa Rica^ies. *Rev. biol. trop* 2014;**62**(3):869-876.
21. Nunes B, Silva RD, Cruz VT, et al. Prevalence and pattern of cognitive impairment in rural and urban populations from Northern Portugal. *BMC Neurology Vol 10 2010, ArtID 42* 2010;**10**.
22. Pilleron S, Clement JP, Ndamba-Bandzouzi B, et al. Is dependent personality disorder associated with mild cognitive impairment and dementia in Central Africa? A result from the EPIDEMCA programme. *International Psychogeriatrics* 2015;**27**(2):279-288.
23. Poddar K, Kant S, Singh A, Singh TB. An epidemiological study of dementia among the habitants of Eastern Uttar Pradesh, India. *Annals of the Indian Academy of Neurology* 2011;**14**(3):164-168.
24. Prince M, Acosta D, Ferri CP, et al. Dementia incidence and mortality in middle-income countries, and associations with indicators of cognitive reserve: A 10/66 Dementia Research Group population-based cohort study. *The Lancet* 2012;**380**(9836):50-58.
25. Raina SK, Raina S, Chander V, et al. Is dementia differentially distributed? A study on the prevalence of dementia in migrant, urban, rural, and tribal elderly population of Himalayan region in Northern India. *North American Journal of Medical Sciences* 2014;**6**(4):172-177.
26. Rodriguez-Agudelo Y, Solis-Vivanco R, Acosta-Castillo I, et al. Neuropsychiatric symptoms in older adulte with and without dementia in urban and rural regions. Results of the 10/66 Dementia Research Group in Mexico. [Spanish]

Sintomas neuropsiquiatricos en adultos mayores con y sin demencia de regiones urbana y rural. Resultados del Grupo de Investigacion en Demencia 10/66 en Mexico. *Revista de Investigacion Clinica* 2011;**63**(4):382-390.
27. Samba H, Guerchet M, Bandzouzi BN, et al. Incidence of dementia among older adults in central africa: First results from the republic of congo in the EPIDEMCA-FU study. *Alzheimer's and Dementia* 2015;**1)**:P221-P222.
28. Sharifi F, Fakhrzadeh H, Varmaghani M, et al. Prevalence of Dementia and Associated Factors among Older Adults in Iran: National Elderly Health Survey (NEHS). *Archives of Iranian Medicine* 2016;**19**(12):838-844.
29. Taylor CA, Greenlund SF, McGuire LC, Lu H, Croft JB. Deaths from Alzheimer's Disease - United States, 1999-2014. *Mmwr* 2017;**Morbidity and mortality weekly report. 66**(20):521-526.
30. Tengku Aizan H, Krishnaswamy S, Siti Suhailah A, Yadollah Abolfathi M. Sociodemographic risk factors and correlates of dementia in older Malaysians. *Dementia and Geriatric Cognitive Disorders* 2010;**30**(6):533-539.
31. Tola-Arribas MA, Yugueros MI, Garea MJ, et al. Prevalence of dementia and subtypes in Valladolid, northwestern Spain: The DEMINVALL Study. *PLoS ONE* 2013;**8**(10).
32. Tornau F, Bruchmann G, Juckel G. Prevalence of psychiatric disorders in the elderly in rural and urban areas in Germany. *International Psychogeriatrics* 2015;**27**:S167.
33. Tripathi M, Vibha D, Gupta P, et al. Risk factors of dementia in North India: A case-control study. *Aging & Mental Health* 2012;**16**(2):228-235.
34. Weden MM, Shih RA, Kabeto MU, Langa KM. Secular trends in dementia and cognitive impairment of U.S. rural and urban older adults. *American Journal of Preventive Medicine* 2018;**54**(2):164-172.
35. Wu YT, Prina AM, Jones A, et al. The Built Environment and Cognitive Disorders: Results From the Cognitive Function and Ageing Study II. *American Journal of Preventive Medicine* 2017;**53**(1):25-32.

36. Yin P, Feng X, Astell-Burt T, et al. Temporal trends and geographic variations in dementia mortality in China between 2006 and 2012: Multilevel evidence from a nationally representative sample. *Alzheimer Disease and Associated Disorders* 2016;**30**(4):348-353.
37. Yuan J, Zhang Z, Wen H, et al. Incidence of dementia and subtypes: A cohort study in four regions in China. *Alzheimer's & Dementia: The Journal of the Alzheimer's Association* 2016;**12**(3):262-271.
38. Zhao Q, Zhou B, Ding D, Guo Q, Hong Z. Prevalence, mortality, and predictive factors on survival of dementia in Shanghai, China. *Alzheimer Disease and Associated Disorders* 2010;**24**(2):151-158.
