## Appendix 6 for "Geographical variation in dementia: systematic review with meta-analysis"

| Study | Country | Total N | Dementia | OR (95% CI) |
|---|---|---|---|---|
| Goodman 2017 | USA | 21,624,228 | 3,110,654 | 0.84 (0.84, 0.85) |
| Chen 2017 | Canada | 2,066,639 | 257,816 | 0.91 (0.90, 0.92) |
| Abner 2016 | USA | 1,209,976 | 82,278 | 0.89 (0.88, 0.91) |
| **High Income Countries: N>1,000,000** | | | | **0.88 (0.84, 0.92)** |
| Astell-Burt 2018 | Australia | 261,669 | 3,046 | 0.63 (0.56, 0.73) |
| Jean 1996 | Canada | 131,667 | 219 | 1.52 (1.21, 1.90) |
| Weden 2018a | USA | 16,386 | 958 | 1.34 (1.10, 1.63) |
| Weden 2018b | USA | 16,311 | 727 | 1.17 (0.92, 1.48) |
| **High Income Countries: N>10,000** | | | | **0.99 (0.80, 1.23)** |
| Koller 2010 | Germany | 9,216 | 1,846 | 0.96 (0.86, 1.08) |
| Wu 2017 | England | 7,505 | 328 | 0.92 (0.64, 1.33) |
| Matthews 2005 | UK | 3,557 | 217 | 0.89 (0.70, 1.13) |
| Hendrie 2018 | USA | 3,276 | 285 | 1.56 (1.21, 2.01) |
| Arslantas 2009 | Turkey | 3,100 | 262 | 4.89 (3.87, 6.19) |
| Lin 1998 | Taiwan | 2,915 | 108 | 1.13 (0.81, 1.59) |
| Ogunniyi 2000b | USA | 2,212 | 65 | 2.49 (1.21, 5.12) |
| Tola-Arribas 2013 | Spain | 2,170 | 184 | 0.99 (0.72, 1.37) |
| Kim 2011 | South Korea | 1,673 | 351 | 1.17 (1.00, 1.38) |
| Yip 1997 | Taiwan | 1,443 | 29 | 1.16 (0.59, 2.25) |
| Goncalves-Pereira 2017 | Portugal | 1,405 | 129 | 0.99 (0.69, 1.42) |
| Nunes 2010 | Portugal | 1,146 | 31 | 2.12 (0.91, 4.96) |
| Azzimondi 1998 | Italy | 727 | 196 | 0.68 (0.52, 0.90) |
| **All High Income Countries** | | | | **1.16 (0.94, 1.42)** |
| Zhang 2006 | China | 34,807 | 1,027 | 1.50 (1.29, 1.75) |
| Yuan 2016 | China | 12,881 | 311 | 1.32 (1.05, 1.66) |
| Jia 2014 | China | 10,276 | 528 | 1.45 (1.22, 1.73) |
| **Low-to-Middle Income Countries: N>10,000** | | | | **1.45 (1.30, 1.60)** |
| Chen 2012b | China | 3,327 | 341 | 2.26 (1.79, 2.86) |
| Tengku Aizan 2010 | Malaysia | 2,980 | 418 | 1.87 (1.52, 2.31) |
| Chen 2012a | China | 2,917 | 210 | 4.27 (3.13, 5.84) |
| Poddar 2011 | India | 2,890 | 146 | 1.49 (0.99, 2.24) |
| Ogunniyi 2000a | Nigeria | 2,494 | 28 | 1.57 (0.44, 5.56) |
| Prince 2012c | China | 2,162 | 161 | 0.93 (0.67, 1.29) |
| Rodriguez 2008c | China | 2,162 | 59 | 0.79 (0.51, 1.23) |
| Prince 2012b | Mexico | 2,003 | 121 | 2.24 (1.51, 3.31) |
| Rodrİguez-Agudelo 2011 | Mexico | 2,003 | 171 | 0.99 (0.72, 1.36) |
| Rodriguez 2008d | India | 1,985 | 17 | 0.88 (0.39, 1.96) |
| Rodriguez 2008b | Mexico | 2,002 | 62 | 0.54 (0.35, 0.84) |
| Prince 2012a | Peru | 1,933 | 69 | 1.36 (0.82, 2.23) |
| Rodriguez 2008a | Peru | 1,931 | 43 | 0.12 (0.04, 0.39) |
| Deng 2018 | China | 1,781 | 195 | 1.56 (0.99, 2.45) |
| Sharifi 2016 | Iran | 1,257 | 99 | 1.88 (1.23, 2.86) |
| Guerchet 2013b | ROC | 1,029 | 63 | 0.85 (0.51, 1.42) |
| Raina 2014 | India | 1,000 | 23 | 0.43 (0.18, 1.05) |
| **Low-to-Middle Income Countries: N>1000** | | | | **1.24 (0.95, 1.63)** |
| Guerchet 2013a | CAR | 973 | 72 | 1.35 (0.83, 2.19) |
| Samba 2015 | ROC | 966 | 23 | 1.78 (0.75, 4.23) |
| Pilleron 2015b | ROC | 912 | 135 | 1.01 (0.60, 1.68) |
| Pilleron 2015a | CAR | 860 | 135 | 1.43 (0.88, 2.32) |
| Khedr 2015 | Egypt | 691 | 35 | 0.44 (0.22, 0.90) |
| Tripathi 2012 | India | 300 | 150 | 0.56 (0.35, 0.90) |
| **All Low-to-Middle Income Countries** | | | | **1.18 (0.94, 1.49)** |
| **All Studies** | | | | **1.18 (1.01, 1.37)** |

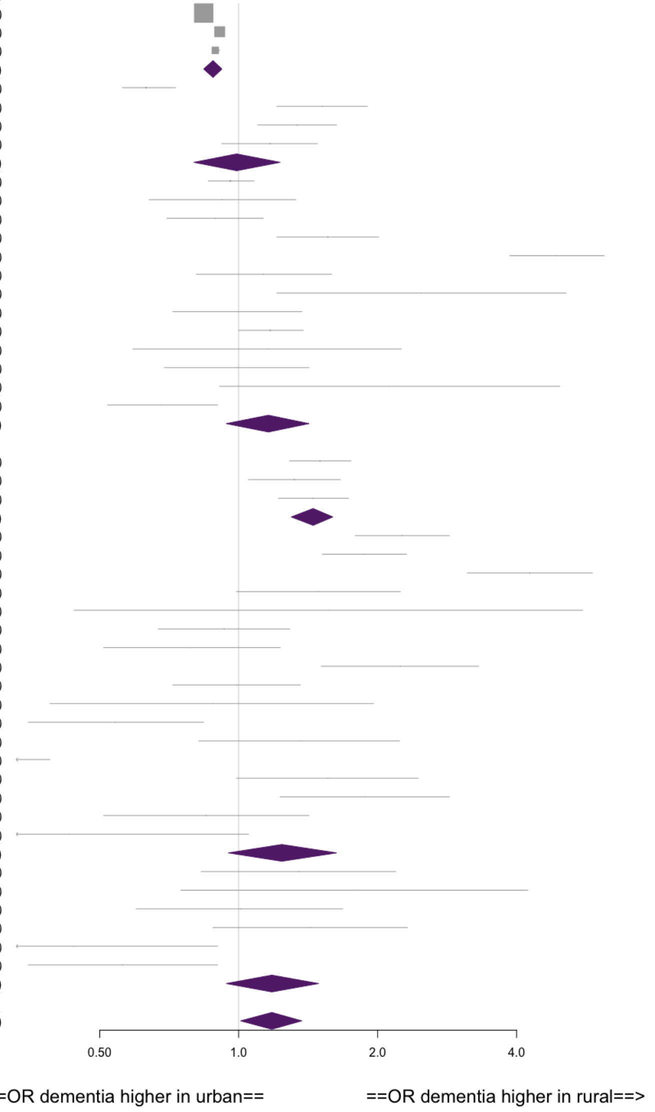


**Appendix 6: Supplementary Figure – Forest plot of all studies in current systematic review and previous 2012 systematic review**
