## Supplementary figures and images for "Geographical variation in dementia: systematic review with meta-analysis"

### Appendix 7

#### Appendix 7

#### Supplementary Figure 3: Risk of Bias assessment by study

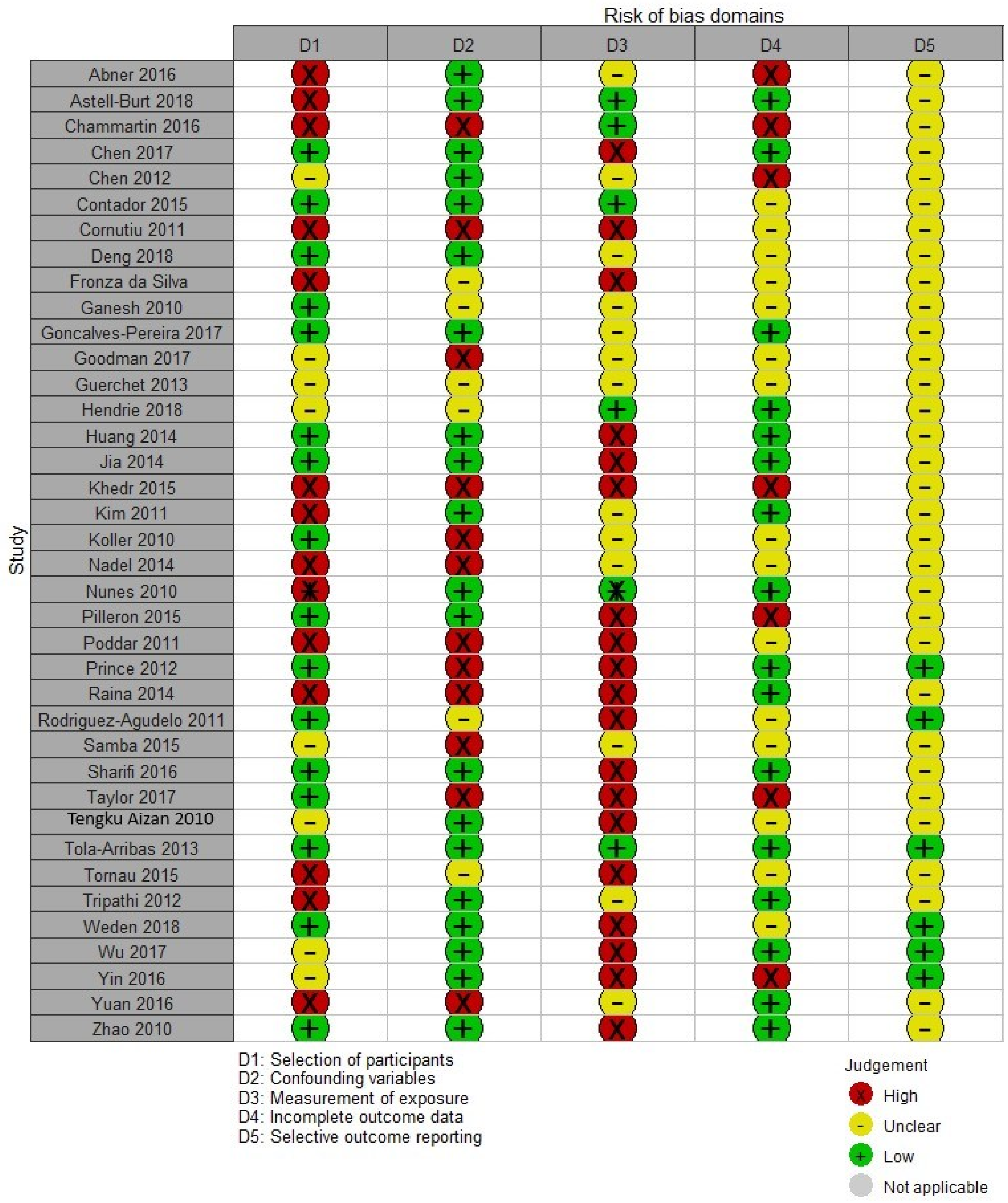
